# Intense *TP53* pathway selection drives clonal evolution from bone marrow failure to leukemia in ERCC6L2 disease

**DOI:** 10.64898/2026.09.16.26363199

**Authors:** SPM Douglas, I Kaaja, I Ikonen, L Langohr, JR Koski, M Hakkarainen, L Katainen, G Barbany, E Hellström-Lindberg, K Jahnukainen, S Kakko, T Siitonen, R Niinimäki, E Pitkänen, U Wartiovaara-Kautto, O Kilpivaara

## Abstract

ERCC6L2 disease (ED) is an inherited bone marrow failure (BMF) syndrome that progresses almost exclusively to erythroid, *TP53*-mutated myeloid malignancy, but the somatic evolution underlying the progression in patients is unknown. We characterized the genetic landscape of 29 ED patients with longitudinal sampling, integrating whole-genome and single-cell RNA sequencing, and compared the findings with Shwachman-Diamond syndrome (SDS), another *TP53* mutation-prone disease, to define somatic evolution across disease stages. ED was defined by early, intense, and recurrent selection of *TP53*-mutant clones: *TP53* mutations were more frequent, more often multiple, and showed steeper age-related expansion than in SDS; all *TP53*-negative ED patients were children. The burden of multiple *TP53* mutations exceeded that of sporadic *TP53*-mutated myeloid neoplasia and was evident before malignancy. Missense mutations dominated the BMF phase, whereas non-missense mutations were seen in patients with malignancy, consistent with biallelic second hits, marking a stage-dependent shift in p53 inactivation. Beyond *TP53*, the landscape was strikingly restricted, with *PPM1D* as the only recurrent driver gene. Notably, despite the DNA-repair function of ERCC6L2, no ED-specific mutational signature emerged; but instead reflected age-associated processes. Malignant progression was accompanied by a complex karyotype, recurrent chromosome 5, 7, and 12p losses enriched in erythroid progenitors, and kataegis, consistent with branched, erythroid-predominant clonal evolution. Our findings establish ED as a paradigm of preleukemic dependency on p53-pathway attenuation. As progression is driven predominantly by p53 and captured longitudinally from the earliest clones, ED offers a model of how *TP53*-mutated leukemia arises and evolves, with relevance beyond a rare syndrome.

## INTRODUCTION

ERCC6L2 disease (ED) is a recessively inherited bone marrow failure syndrome (IBMFS) with a propensity for *TP53* mutations^1–3^. ED-associated bone marrow failure (ED-BMF) progresses to myeloid malignancy almost exclusively with erythroid features^1^ with a median onset in the 30s^2^. Given the absence of effective therapies for *TP53*-mutated myeloid malignancies, the prognosis of ED remains poor. Under current classification these erythroid-predominant malignancies are considered MDS or AML with mutated *TP53*, yet only 10% of *TP53*-mutated AMLs are erythroid. *TP53*-mutated AMLs are more prevalent if preceded by another hematological malignancy (HM) or anti-cancer therapy^4^. More broadly, *TP53*-mutated myeloid malignancies are strongly associated with complex karyotype^5^ and other structural genomic rearrangements, including chromothripsis^6^, a catastrophic event involving extensive chromosome fragmentation and reassembly.

ERCC6L2 has established roles in DNA repair, centromere stability, and immunoglobulin class-switch recombination, with putative mitochondrial involvement ^7–11^, and was recently implicated in DNA end resection during repair of staggered-end double-strand breaks ^12,13^; however, mechanistic understanding derives almost exclusively from non-patient systems. Most recently, murine and *in vitro* models demonstrated that loss of Ercc6l2 induces replication stress, whereas somatic *Trp53* mutations restore cellular fitness at the cost of increased leukemogenic potential^14^. These findings predict positive selection of *TP53*-mutant clones as a mechanism to relieve ERCC6L2-driven stress, but the genomic dynamics of this process in patients remain uncharacterized.

Recent single-cell studies of primary ED samples showed early emergence of *TP53*-mutant progenitor clones and transcriptional parallels with Shwachman-Diamond syndrome (SDS), another IBMFS predisposing to *TP53* mutagenesis and sharing erythroid stress programs^3^; but unlike ED, SDS is not associated with erythroid-type myeloid malignancy^15^.

Here, we outline the somatic genetic landscape across 29 ED patients, most with longitudinal sampling, and compare disease progression with SDS (n = 18). Using whole-genome sequencing (WGS) integrated with single-cell RNA sequencing (scRNA-seq) to map copy-number alterations (CNA) onto bone marrow (BM) cell populations, we define the timing and genomic consequences of p53 pathway disruption, clonal selection, and malignant transformation. This first genome-wide, patient-derived characterization of somatic evolution in ED allows direct assessment of whether mechanisms proposed in cell-line and animal models are reflected in patients during progression from ED-BMF to leukemia.

## MATERIALS AND METHODS

### Patients

We collected clinical data, including clinical panel sequencing and karyotyping, and BM or blood and normal tissue samples (Table S1, Supplementary Information) from all available ED patients (n = 29; 28 from Finland and one from Sweden) and SDS patients (n=18) from Finland^2,16^. Clinical history and detailed information on disease characteristics were gathered from the Finnish Hematological Registry and Biobank (FHRB) and patient records. Patients are summarized in Table S2 with details about clinical data in Supplementary Information.

### Whole-genome sequencing and secondary data analysis

We conducted whole-genome sequencing (WGS) on 27 ED BM/blood samples (from 21 patients) and 4 SDS patients and corresponding normal tissue samples from 18 ED patients and 4 SDS patients (Table S1). WGS libraries were prepared using standard protocols and sequenced as 2 × 150 bp reads on a NovaSeq 6000 (Illumina, San Diego, CA). Secondary analysis was performed with Illumina DRAGEN Bio-IT Platform (v4.4.6). Details and coverage metrics are provided in Supplementary Information.

### Somatic mutations

Somatic non-synonymous coding and splice-site variants were filtered to include only high-confidence variants (Supplementary Information). Potential somatic driver mutations were identified with Cancer Genome Interpreter (CGI)^17^ (v23.12.2), (https://www.cancergenomeinterpreter.org/), using MDS as cancer type. All genes with somatic mutations in three or more ED patients with paired tumor-normal WGS data were also assessed.

### Copy-number alterations and structural variants

DRAGEN, and Sequenza (v.3.0.0)^18^, were used for calling somatic CNAs. Chromosome-level aberrations were assessed by integrating clinical karyotyping data (where available), DRAGEN CNA calls, segmentation and B-allele frequency (BAF) calls and Sequenza CNA results. Dragen structural variant (SV) calls that had passed filtering, and not flagged as imprecise, were further filtered (Supplementary Information) to include only high-confidence calls.

Filtered somatic CNAs called by DRAGEN and high-confidence SVs, were analyzed with CGI using MDS as cancer type. All large CNAs and SVs were visually inspected with Integrative Genomics Viewer (IGV) (v.2.14.0), with only clear alterations reported.

### Biallelic *TP53* inactivation status

Biallelic *TP53* inactivation was inferred from *TP53* mutation variant allele fraction (VAF), CNA and copy-neutral loss-of-heterozygosity (CN-LOH) status at the *TP53* locus and estimated sample purity. Cases were classified as confirmed biallelic when a *TP53* mutation co-occurred with 17p/*TP53* locus deletion or CN-LOH. Cases with multiple *TP53* mutations were considered biallelic when two mutations were in trans and their combined VAF exceeded 50%, with the observed VAFs being consistent with the estimated sample purity. Cases with two *TP53* mutations with combined VAF>50% or a single mutation with VAF >50% were considered likely biallelic. Samples without WGS data, phasing of variants and smaller VAFs were considered not assessable.

### Chromothripsis

Presence of chromothripsis was analyzed with Shatterseek^19^ (v.1.1) from DRAGEN CNA calls and manually curated SVs according to the recommended cut-off values^19^.

### Mutational signatures and clustered somatic hypermutation

We analyzed mutational signatures and somatic hypermutation patterns in somatic samples with a matching normal sample using SigProfiler^20^ and SigProfilerClusters^21^.

Detected kataegis variants were visually inspected and suspicious clusters were discarded.

### Differential expression in pseudobulk from scRNA-seq data

Pseudobulk were generated based on published ED scRNA-seq data^3^. Pseudobulk per sample were generated by summing the gene expression per gene and donor, by adapting the pseudobulk code from the Single-cell best practices -book^22^. Differential expression was analyzed with DESeq2 (1.52.0)^23^ with default parameters.

### Copy-number alterations from scRNA-seq data

To identify CNAs using scRNA-seq data, we used inferCNVpy (v.0.5.0) as described before^3^.

### Sporadic comparator dataset

*TP53* mutation-count distribution per-patient for sporadic myeloid neoplasia was derived from the MSKCC 2020 myeloid neoplasm dataset^24–26^, retrieved via the cBioPortal^27,28^ REST API. Patients of all AML and MDS subtypes were included in the analysis. Silent/synonymous, intronic, and untranslated-region variants were excluded to approximate a pathogenic/likely-pathogenic filter, and large deletions and copy-number events were excluded from both the sporadic and ED counts, ensuring comparability (n = 351 *TP53*-mutated patients). Replicate samples from the same patient were deduplicated. Claude (Opus 4.8; Anthropic, San Francisco, CA, USA) was used for data collection and analysis.

### Statistical analyses

Continuous variables were compared between ED and SDS using the Wilcoxon rank-sum test, and categorical variables using Fisher’s exact test. Logistic regression was used to assess associations after adjustment for age. Poisson regression was used to compare mutation counts between groups. Fractional logistic regression with a quasibinomial distribution and logit link was used to assess the association between maximum *TP53* variant allele frequency (VAF) and age separately in ED and SDS. Odds ratios (ORs) with 95% confidence intervals (CIs) were reported for regression analyses. For contingency-table analyses, ORs with 95% CIs were calculated using the Haldane–Anscombe correction when applicable. Statistical significance was defined as *P* < 0.05. Statistical analyses were performed in R (v.4.4.2).

## RESULTS

### Patient characteristics

Median age at last follow up was 37 (range 6-65) for ED patients, and 24 (10-70) for SDS patients. After adjusting for age, ED remained associated with higher odds of HM (OR 6.8, 95% CI 1.45–42.9, *P* = 0.023) and allogeneic hematopoietic stem cell transplantation (HSCT) (OR 6.8, 95% CI 1.81–31.03, *P* = 0.007) than SDS.

Among the 18 ED patients with HM, nine had MDS, five erythroid AML, three MDS with excess blasts (MDS/AML), and one monocytic AML. All three SDS patients with HM had MDS. The median age among patients with HM was similar between groups: 36.9 years (range, 14–65) for ED and 35.2 years (range, 23–70) for SDS. Patient characteristics are summarized in Table 1. Longitudinal data were available for 22/29 ED patients (75.9%) and 17/18 SDS patients (94.4%).

**Table 1.** Patient characteristics for ED and SDS patients. Characteristics are summarized at the latest follow-up for treatment-naïve patients or the last timepoint before potential treatment or allogeneic hematopoietic stem cell transplantation (HSCT). Follow-up time is calculated from time with sampling. AML, acute myeloid leukemia; BM, bone marrow; BMF, bone marrow failure; MDS, myelodysplastic syndrome.

|  | **ED** | **SDS** | **P (age-adjusted)** |
| --- | --- | --- | --- |
| All patients, n | 29 | 18 |  |
| Age, median (range), years (last timepoint) | 36.7 (6-65) | 24.1 (10-70) | 0.031 |
| Sex, n (%) |  |  |  |
| Male | 15 (51.7%) | 13 (72.2%) | 0.23 |
| Female | 14 (48.3%) | 5 (27.8%) |  |
| Non-malignant, n (%) (last timepoint) | 11 (38.0%) | 15 (83.3%) |  |
| Age, median (range) years | 20.6 (6-65) | 22.8 (6-46) |  |
| Sex, n (%) |  |  |  |
| Male | 6 (54.5%) | 10 (66.7%) |  |
| Female | 5 (45.5%) | 5 (33.3%) |  |
| BMF | 10 (90.9%) | 15 (100%) |  |
| Normocellular BM | 1 (9.1%) | 0 |  |
| Malignant, n (%) (last timepoint) | 18 (62.1%) | 3 (16.7%) | 0.0029 (0.023) |
| Age, median (range) years | 36.9 (14-56) | 35.2 (23-70) | 0.88 |
| Sex, n (%) |  |  |  |
| Male | 9 (50%) | 3 (100%) |  |
| Female | 9 (50%) | 0 |  |
| MDS | 9 (50%) | 3 (100%) |  |
| MDS/AML | 3 (16.7%) | 0 |  |
| erythroid AML | 5 (27.8%) | 0 |  |
| monocytic AML | 1 (5.6%) | 0 |  |
| HSCT, n (%) | 19 (65.5%) | 4 (22.2%) | 0.0065 (0.007) |
| Alive, n (%) | 17 (58.6%) | 17 (94.4%) | 0.0084 (0.064) |
| Follow up time, median (range) years | 2.6 (0-14.7) | 4.0 (0.3-16.9) |  |

### ED patients show strong selection for *TP53* pathway mutations

*TP53* mutations were detected in 26/29 (90%) ED patients at last follow-up and 8/18 (44%) of SDS patients (Fig. 1, Fig. 2A, Table S3) and were significantly more frequent in ED (*P* < 0.002; OR 10.17, 95% CI 2.00–71.92) (Fig. 2A). The *TP53* mutation frequency in our SDS cohort was comparable to previous reports on SDS^29^. ED patients were more likely to harbor multiple *TP53* mutations (*P* = 0.006; OR 6.4, 95% CI 1.49–33.93) (Fig. 2B) and carried more *TP53* mutations per patient (*P = 7.58 × 10*⁻□) (Fig. 1B and 2C) than SDS patients. In an age-adjusted Poisson model, *TP53* mutation counts were approximately 2.4-fold higher in ED than in SDS (*P =* 0.003) (Fig. 2D).

**Fig. 1.**
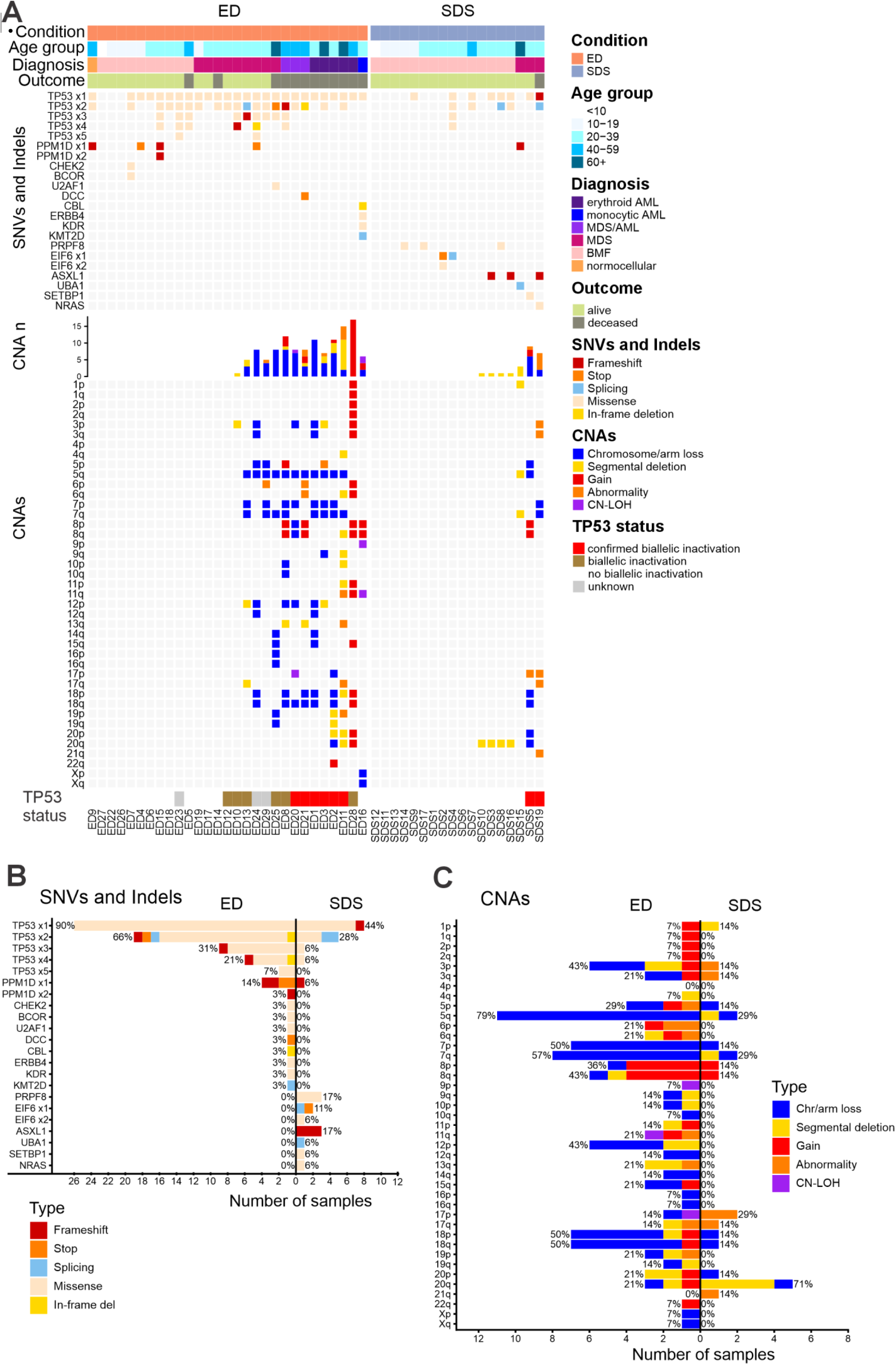
Landscape of somatic alterations in ED and SDS. **A** Somatic single nucleotide variants (SNVs), small insertions and deletions (indels) and chromosome-level copy-number alterations (CNAs) at last follow-up (or last timepoint before hematopoietic stem cell transplantation/treatment). Variants predicted as drivers or reported by clinical panel sequencing are shown. **B** Number and percentage of ED and SDS patients with somatic SNVs and indels in potential driver genes by alteration type. **C** Number and percentage of each CNA type in ED and SDS patients with CNAs. AML, acute myeloid leukemia; BMF, bone marrow failure; chr, chromosome; CN-LOH, copy-neutral loss of heterozygosity MDS, myelodysplastic syndrome.

**Fig. 2.**
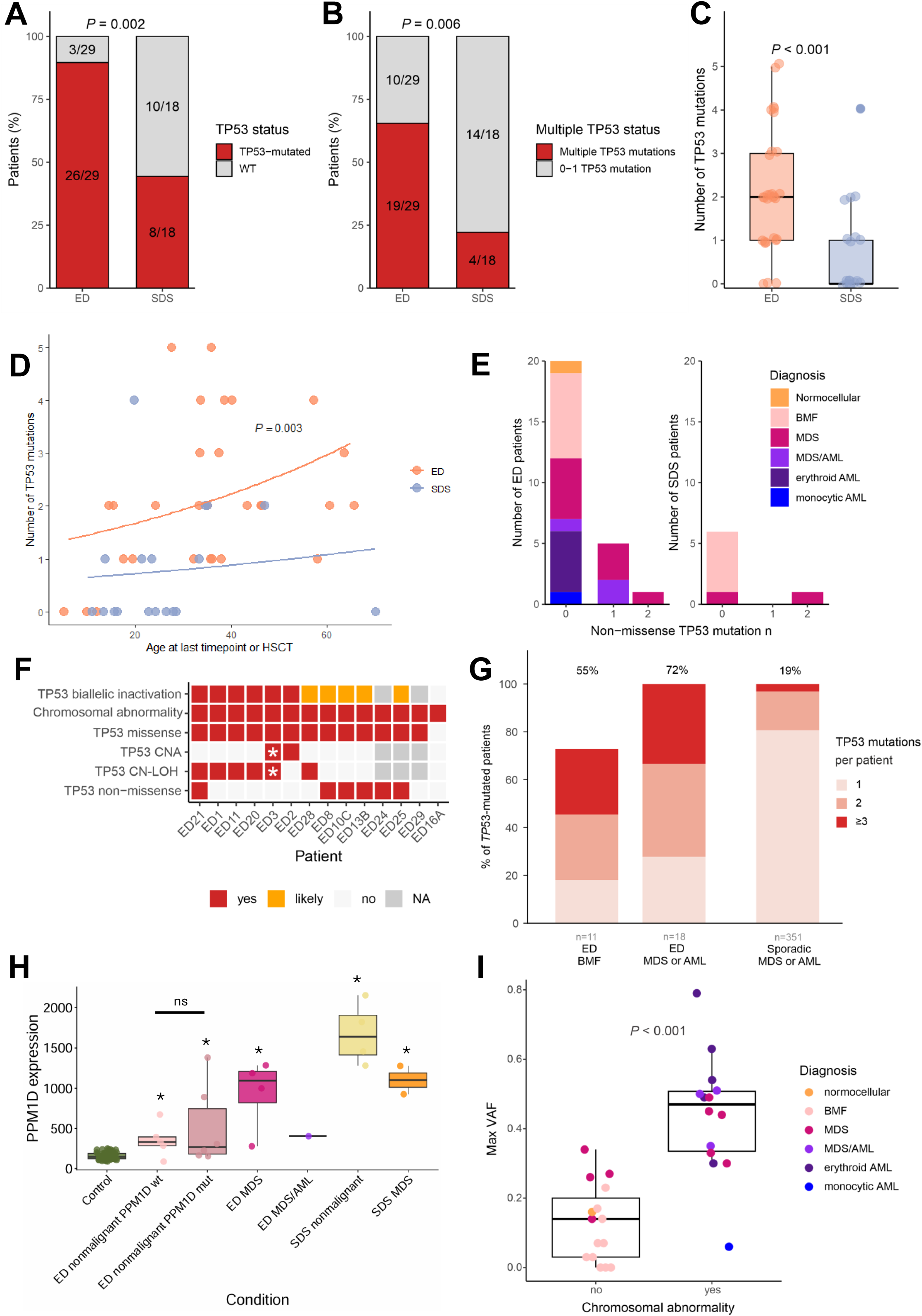
*TP53* pathway mutational burden in ED and SDS. **A** Proportion of patients with at least one somatic *TP53* mutation compared to wild-type patients at the latest follow-up or last pre-treatment/pre-HSCT timepoint in ED and SDS. **B** Proportion of patients with ≥2 *TP53* mutations compared to those with 0-1 *TP53* mutations in ED and SDS. **C** Number of *TP53* mutations per patient in ED and SDS. Boxes show median and interquartile range; points indicate individual patients. **D** Number of *TP53* mutations per patient plotted against age at the latest follow-up or last pre-treatment/pre-HSCT timepoint in ED and SDS. Lines indicate fitted Poisson regression curves by condition. The *P* value denotes the age-adjusted difference in *TP53* mutation count between ED and SDS. **E** Stacked bar plots show the number of patients with 0, 1, or 2 non-missense *TP53* mutations in ED (left) and SDS (right), stratified by diagnosis at the latest follow-up or last pre-treatment/pre-HSCT timepoint. **F** *TP53* alteration status in ED patients with chromosomal abnormalities. Timepoint with WGS data was used if available. \**TP53* gain-associated loss of heterozygosity. **G** Proportion of *TP53*-mutated patients carrying one (grey), two (amber), or three or more (dark red) pathogenic/likely-pathogenic *TP53* mutations, in ED at the bone marrow failure (BMF) stage and at the malignant (MDS or AML) stage, compared with sporadic MDS or AML. Percentages above each bar indicate the proportion of patients with ≥2 mutations within the diagnosis group. **H** *PPM1D* expression in the bone marrow samples by diagnosis groups in ED and SDS, compared to healthy controls. Points represent individual samples. *Differentially expressed (Adjusted *P* < 0.05) vs. control. **I** Comparison of largest *TP53* VAF in ED samples without or with chromosomal abnormalities. Boxes show median and interquartile range; points represent individual samples and are colored by diagnosis. BMF, bone marrow failure; HSCT, allogeneic hematopoietic stem cell transplantation; MDS/AML, myelodysplastic syndrome/acute myeloid leukemia; NA, not assessable; VAF, variant allele fraction.

The age distribution further supported early *TP53*-mutant clonal selection in ED: all *TP53*-mutation-negative ED patients were younger than 13 years (vs. under 71 for SDS), and the median age of *TP53*-negative patients was lower in ED than SDS (10.3 vs 25.1 years, *P* = 0.003), including the last pre-mutation samples for patients who later became *TP53*-positive. Maximum *TP53* VAF increased significantly with age in ED (OR 1.050 per year, 95% CI 1.03–1.07, *P =* 1.49 × 10⁻□), whereas no significant age-associated increase was observed in SDS (OR 1.017 per year, 95% CI 0.95–1.09, *P* = 0.653). The evolution of *TP53* and other somatic mutation VAFs over time in each ED patient is shown in Supplementary Fig. 2-5.

The *TP53* mutations clustered mostly in the hotspot exons 5-9 (Supplementary Fig. 6A). Non-missense *TP53* mutations (n=7, in six patients) in ED were seen only in MDS patients with chromosomal abnormalities (Fig. 2E-F), and had lower VAFs than the dominant *TP53*mutation, suggesting later acquisition in clonal evolution. In three samples, the non-missense mutation was clearly on the opposite allele from the mutation with the largest VAF (*in trans*) (Table S4). Moreover, three of the non-missense mutations were in samples with no identified *TP53* loss through CNA or CN-LOH (Supplementary Fig. 7). In all cases, the non-missense mutation VAF was compatible with estimated purity of the sample or aberrant fraction in karyotyping, strongly suggesting a mechanism for biallelic p53 inactivation in ED (Fig. 2F, Table S4). In SDS, non-missense *TP53* mutations occurred in two patients (Fig. 2E, Table S3).

ED patients carried substantially more pathogenic/likely-pathogenic *TP53* mutations per patient than observed in sporadic *TP53*-mutated myeloid neoplasia (Fig. 2G). Among *TP53*-mutated ED patients with MDS or AML, 72% (13/18) carried two or more distinct *TP53* mutations, compared with 19% (68/351) of sporadic MDS or AML patients (OR 10.8, 95% CI 3.6–28.3, *P =* 3.96 × 10⁻ ) (Fig. 2G). The tendency to harbor multiple *TP53* mutations was already present before malignant transformation: 55% (6/11) of ED-BMF patients carried two or more *TP53* mutations, exceeding the rate in sporadic MDS or AML (OR 5.0, 95% CI 1.5–15.7, *P* = 0.012) (Fig. 2G). High-multiplicity clones were particularly enriched in ED: three or more *TP53* mutations were present in 33% of ED MDS or AML and 27% of ED-BMF patients, but in only 3% of sporadic patients (Fig. 2G). The mean *TP53* mutation count was 2.3 in ED MDS or AML and 1.9 in ED-BMF, versus 1.3 in sporadic MDS or AML.

Together, these findings indicate that ED evolution is highly dependent on *TP53* mutagenesis, with earlier and stronger selection than in SDS and a greater burden of multiple *TP53* mutations than in sporadic *TP53*-mutated MDS or AML. Furthermore, non-missense *TP53* mutations were associated with malignant progression in ED.

### *PPM1D* mutations represent additional p53 pathway adaptation in ED

The only other recurrent possible driver mutations in ED occurred in *PPM1D* (4 patients, up to 2 mutations per patient, Fig. 1A-B, Table S3). All were truncating exon 6 mutations (Supplementary Fig. 6B), a recurrent gain-of-function pattern that stabilizes PPM1D and attenuates p53-mediated DNA-damage response^30^. *PPM1D* mutations were detected mostly before malignancy in ED: three patients had one or more *PPM1D* mutations before HM, whereas only one SDS patient with MDS, aged over 60 years, carried a *PPM1D* mutation. In a Poisson regression model without interaction, combined p53 pathway (*TP53* and *PPM1D)* mutation count increased with age (β = 0.0148 per year, *P* = 0.031) and was significantly higher in ED than in SDS (β = −0.892 for SDS vs ED, *P* = 0.002) (Supplementary Fig. 6C), corresponding to an approximately 2.4-fold higher burden in ED. Although *PPM1D* mutations are common in patients after genotoxic therapy^30^, three of the four ED patients with *PPM1D* mutations were treatment-naïve.

*PPM1D* expression was increased in ED cases with *PPM1D* mutations, but also in mutation-negative ED-BMF and MDS cases and in non-malignant SDS samples and rose over time in serial scRNA-seq samples independent of mutation status (Fig. 2H, Supplementary Fig. 6D). However, two ED-BMF patients with high PPM1D VAFs (up to 35%) did not develop HM during follow-up, whereas ED24 had only a minor PPM1D clone (2% VAF) despite MDS (Supplementary Fig. 5, Table S3).

These findings suggest that *PPM1D* mutations and increased *PPM1D* expression reflect additional attenuation of p53-mediated stress responses in ED, but that *PPM1D*-mutant clone expansion alone is insufficient to drive malignant transformation.

### Somatic driver mutations are more heterogeneous in SDS than ED

To determine whether ED and SDS differ in the broader spectrum of somatic drivers beyond *TP53*-pathway lesions, we assessed other putative oncogenic and recurrent non-oncogenic mutations across disease stages.

Four ED patients had additional possible driver mutations (Fig.1A). ED7, a child with ED-BMF, had mutations in *CHEK2* and *BCOR* (VAFs 9% and 21%), alongside a small *TP53* clone (VAF 2%) (Supplementary Fig. 2, Table S3). They were originally classified as variant of uncertain significance (VUS) by a clinical geneticist, but the *CHEK2* variant was suggested as a potential driver by CGI. ED25 and ED21 had predicted drivers in *U2AF1* (VAF 16%) and *DCC* (VAF 7%), both likely subclonal to co-occurring *TP53* mutations. The remaining possibly oncogenic mutations were confined to ED16, the rare non-erythroid (monocytic) leukemia, which carried four possible drivers (*CBL, ERBB4, KDR, KMT2D*) with only a small *TP53* clone (Fig. 1A, Supplementary Fig. 4, Table S3).

SDS patients had recurrent mutations in *PRPF8* (3/18, 17%), *ASXL1* (3/18, 17%) and *EIF6* (2/18, 11%), most already present in the BMF phase (Fig.1A, Table S3). Single patients harbored *UBA1*, *SETBP1* and *NRAS* mutations in the MDS phase. *EIF6* and *PRPF8* were classified as non-oncogenic by CGI; *EIF6* commonly rescues the ribosome defect in SDS and *PRPF8*, mutated in ∼12% of SDS patients, is not associated with HM^29^. In contrast to SDS, we did not find any recurrently mutated non-oncogenic gene in ED patients suggestive of a compensatory mechanism. Also, in *TP53-*mutated SDS patients with MDS, the other driver mutations had large VAFs (*SETBP1* with 34% VAF in SDS5; *ASXL1* with 37% VAF in SDS19, Table S3) suggesting substantial involvement in disease pathogenesis.

Together, these findings propose that somatic evolution outside the p53 pathway is limited in ED and mainly associated with malignant-stage or atypical cases, whereas SDS shows broader clonal diversity, including recurrent disease-adaptive mutations and additional high-VAF drivers in MDS.

### ED patients show recurrent chromosomal abnormality pattern

To determine the frequency, distribution, and recurrence of copy-number changes, we evaluated chromosomal abnormalities across disease stages. ED-BMF patients did not harbor any somatic CNAs (Supplementary Fig. 8-10). In contrast, 14 ED patients with malignancy had at least one chromosomal abnormality, and 13 had complex karyotype defined as three or more independent chromosomal alterations (Fig. 1A, Supplementary Fig. 10). The most common alterations were −5q (79%) and chr7 loss (7q loss in 57%, 7p loss in 50%, monosomy 7 in 43%), followed by chr18 abnormalities (50%), 12p loss (43%), abnormal 3p (43%), and chr8 gain (29%) (Fig. 1C, Table S5). In ED13, scRNA-seq identified cells with −5q, −7, −12p and −17q, missed by clinical karyotyping. In SDS, 7/18 (39%) patients had chromosomal abnormalities, most commonly del(20q) (71%), a known compensatory mechanism in SDS, involving *EIF6*^31^. Additionally, partial or complete loss of 5q and 7q were seen in two patients (29%), as well as abnormality of 17p.

In ED, chromosomal abnormalities were strongly associated with higher *TP53* VAF (*P =* 6.39 × 10⁻□) (Fig. 2I). Median VAF of the biggest *TP53* clone was 14% in patients without chromosomal abnormalities and 49% for *TP53*-driven chromosomally abnormal samples (i.e., excluding ED16).

Regions recurrently deleted in ED comprised broad segments on chromosomes 5q, 7, 12p, 3p, and 13q containing multiple tumor suppressor and myeloid disease genes, while CGI prioritized a narrower set of candidate drivers, including *APC, IKZF1, CDKN1B, BAP1, PBRM1*, and *RB1* (Fig. 3A-E, Tables S6-S7).

**Fig. 3.**
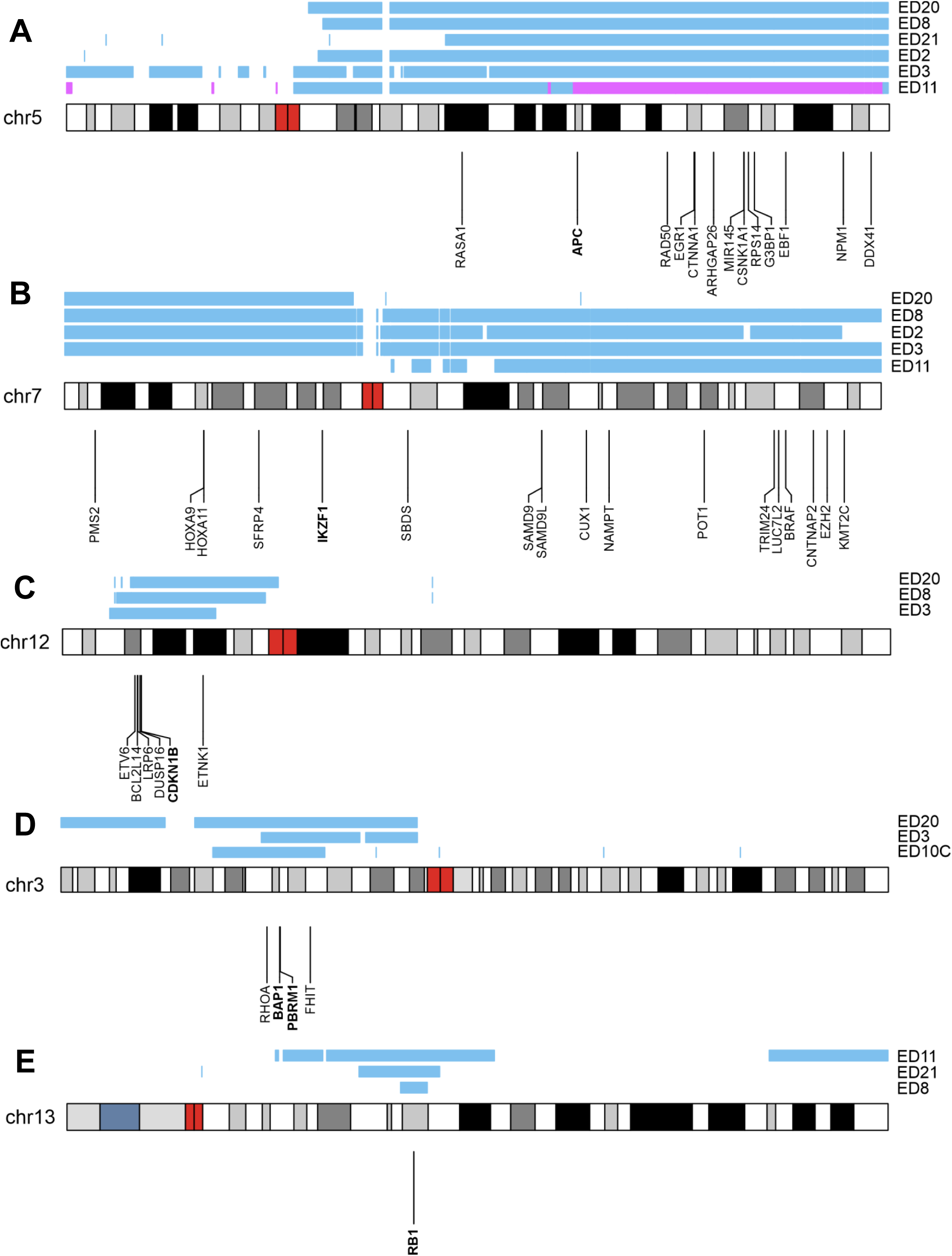
Common deleted regions in ED patients and Cancer Gene Census tumor suppressor genes, myeloid panel genes and other previously suggested driver genes within the regions. **A** Chromosome 5 losses (blue) or copy-neutral loss of heterozygosity (purple) in six ED patients. Shared area covers most of chr5q. **B** Chromosome 7 losses in five patients cover either the whole chromosome or p or q arm only. **C** A deletion in chromosome 12p, covering *ETV6, BCL2L14, LRP6, DUSP16, CDKN1B* and *ETINK1* in three ED patients. **D** In chromosome 3p, the shared deleted area covers *RHOA, BAP1, PBRM1* and *FHIT* in three patients. **E** Chromosome 13q common deleted area covers only *RB1* in all three patients. Genes suggested as drivers by Cancer Genome Interpreter are written in bold.

Integration of longitudinal cytogenetic, WGS, and scRNA-seq data revealed branched patterns of somatic cytogenomic abnormalities in ED (Fig. 4). Recurrent chr5 abnormalities appeared early in at least six patients, whereas chromosome 7 and 12p losses were often within the main clone, while del(3p) and −18 were mostly subclonal and diminished after treatment, consistent with ongoing clonal remodeling (Fig. 4).

**Fig. 4.**
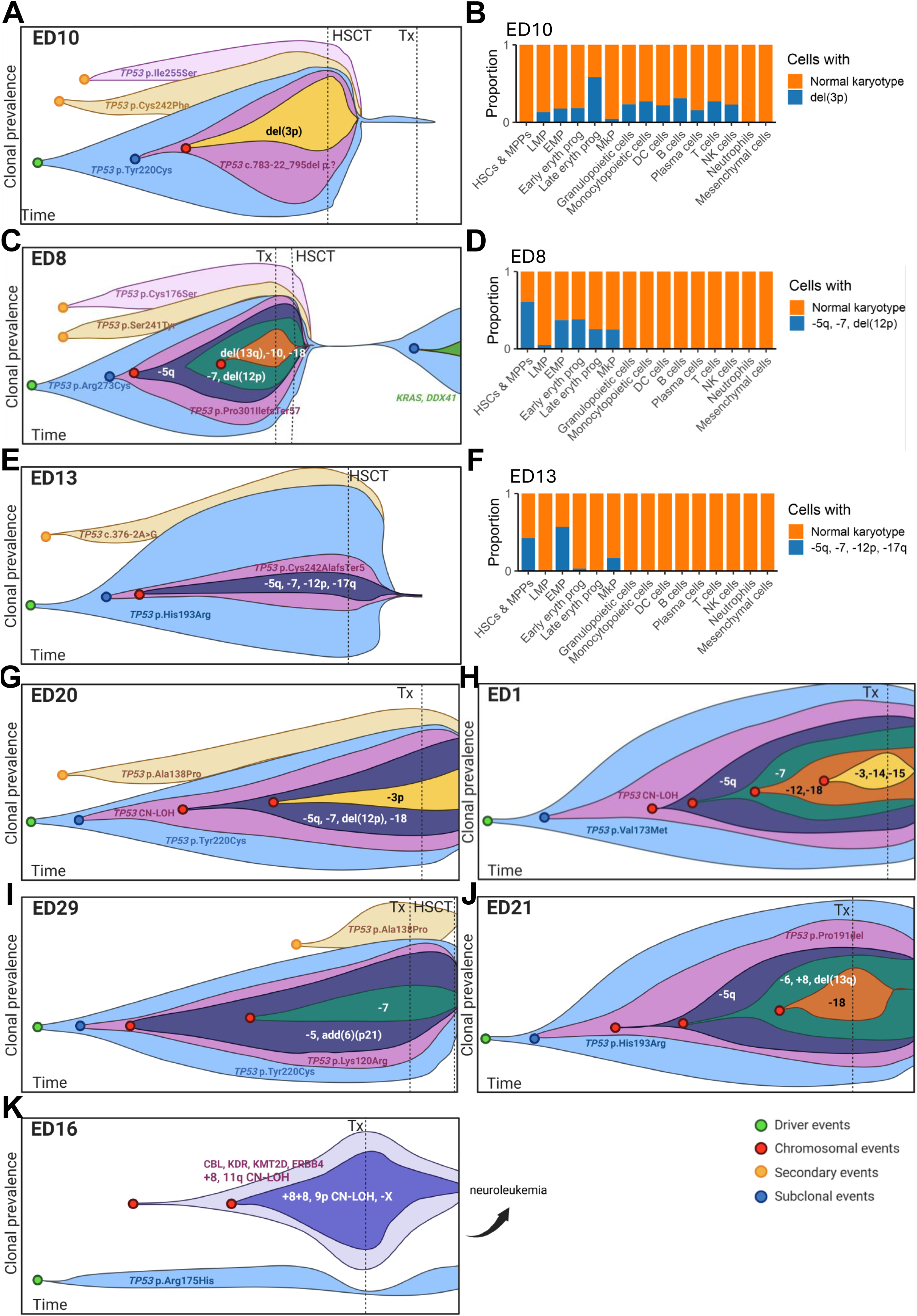
Clonal evolution in ED. **A,C,E,G-K** Illustration of clonal evolution and the likely order of events during disease evolution. Clone sizes and temporal spacing are schematic and not quantitative in the illustration. Loss of 5q is an early aberration followed by or together with –7 (C,E, G-J), often also with del(12p) (C, E, G-H), del(13q) (C, J), -18 and/or del(3p) (C, G, H, J). Aberrations in chromosomes 13 and 18 may disappear after treatment, but -5, -7 and del(12p) seemed resistant to treatments (C, G-H, J). **B,D,F** Copy-number alterations inferred in different cell types from scRNA-seq data. **B** ED10 developed del(3p), seen in most cell types, but enriched in early erythroid progenitors (EEP). **D** In E8 -5, -7, del(12p) were identified mostly in erythroid lineage and megakaryocyte progenitors (MkP) and a smaller clone with del(13q) and -18 that disappeared after treatment was seen at low levels in granulopoietic and monocytopoietic cells in the myeloid lineage. **F** In E13, a small clone with -5q, -7(q), -12p and -17q alterations was mostly detected in erythro-myeloid progenitors (EMP), MkP, hematopoietic stem cells and multipotent progenitors (HSC & MPP) and some in EEP. **K** In ED16, chr8 pentasomy arose in a clone with chr8 trisomy, along with somatic driver mutations and CN-LOH regions. Small *TP53*-mutated clone was not detected at all timepoints. The patient succumbed to neuroleukemia. Del, seqmental deletion; NK, natural killer; LMP, lympho-myeloid progenitor.

Single-cell CNA inference in the two −5/−7/−12p cases (ED8, ED13) localized the lesions mainly to erythroid and megakaryocyte progenitors (Fig. 4, Supplementary Fig. 11). In ED8, +8, −10, del(13q), and −18 disappeared after treatment, with traces of del(13q) and −18 persisting at low levels in myeloid cells, suggesting a clone separate from the treatment-resistant erythroid precursors (Fig. 4, Supplementary Fig. 11). ED10 had an isolated 3p deletion across most cell types but also showed enrichment in early erythroid progenitors (Fig. 4, Supplementary Fig. 12). In SDS5 −5, +8 and −18 spanned the erythroid lineage, but were present at a small proportion of most cell types (Supplementary Fig. 13).

Two cases stood out from the ED cohort: patient ED28, who developed erythroid AML despite a hyperdiploid rather than loss-dominated karyotype, and ED16, whose disease evolved through chr8 trisomy and pentasomy to monocytic AML, likely driven by *MYC* overexpression and not *TP53* (Supplementary Fig. 4, Fig. 4K). Pentasomy of chr8 has been reported before in monocytic leukemia^32^.

Together, these findings show that ED-associated disease progression is marked by the accumulation of multiple chromosomal abnormalities, resulting in complex karyotypes, with −5q and chromosome 7 loss emerging early, followed by −12p, −13q, −18 and −3p as later events with malignant recurrent −5/−7/12p patterns frequently enriched in erythroid-lineage compartments.

### Somatic CNA events and structural variants reflect large-scale genomic instability in advanced ED

Given the role of ERCC6L2 in maintaining genome stability, we examined CNA and SV events as potential drivers of disease progression.

Predicted driver CNAs in ED were concentrated in patients with chromosomal abnormalities and largely reflected broad chromosomal gains, losses, and CN-LOH rather than focal events (Table S8). High-confidence somatic structural variants were absent in BMF and MDS samples but accumulated in MDS/AML and erythroid AML (Supplementary Fig. 14A,7-8). The most common alterations in the *TP53*-driven MDS-AML or AML were inter- and intrachromosomal rearrangements, including inversion-like events, and deletions (Supplementary Fig. 14B). The complex rearrangements in ED patients occurred on chromosomes affected by CNAs, and no recurrent breakpoint-targeted driver genes were identified (Fig. 5, Table S9). Breakpoint homology analysis showed no single dominant repair signature (data not shown), suggesting that multiple DNA break-repair mechanisms contribute to SV formation.

**Fig. 5.**
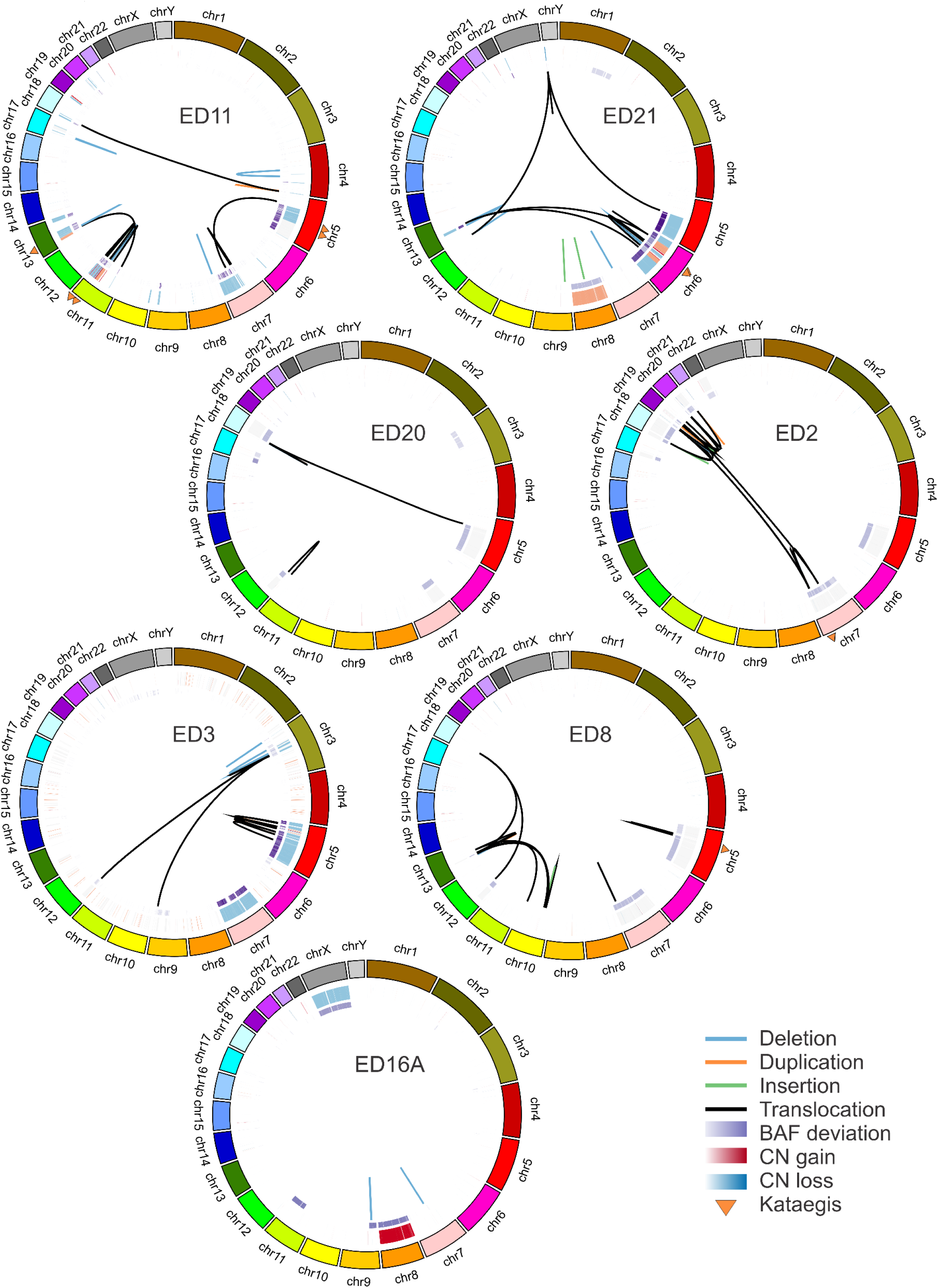
Somatic large-scale aberrations in ED MDS/AML and AML samples. Circos plots show copy number gains (red) and losses (blue), B-allele frequency deviation from 0.5 (purple), structural variants (SVs) (deletion; blue link, duplication; orange, insertion; green, translocation; black) and clustered point mutations. SVs and kataegis cluster with large copy number loss segments in ED patients with *TP53*-driven disease, while ED16A shows different chromosomal abnormalities, few SVs and no kataegis. AML, acute myeloid leukemia; MDS/AML, myelodysplastic syndrome with excess blasts.

To further characterize the complex rearrangements, we assessed chromothripsis. No sample met strict criteria for high-confidence chromothripsis; however, 4/6 samples with complex rearrangements showed chromothripsis-like features, including clustered structural variants and oscillating copy-number states, all occurring in erythroid-type MDS/AML or AML (Table S10).

Our findings indicate that CNA and SV events in ED primarily reflect late, large-scale chromosomal instability in advanced *TP53*-driven disease rather than recurrent focal targeting of individual genes.

### Mutational signatures in ED are dominated by age-associated processes

As ERCC6L2 functions in DNA repair, we investigated whether ED showed distinct mutational processes compared with SDS, that affects ribosome biogenesis. Across WGS samples, most single-base substitutions (SBS) were attributed to the clock-like signatures^33,34^ SBS1 and SBS5 (Fig. 6A,C-D), and no ED-specific SBS signature was identified. Instead, the SNV burden in ED was consistent with age-associated endogenous mutagenesis previously described in normal hematopoiesis and in pediatric AML/HSPC comparisons^35,36^ (Supplementary information). Small insertion and deletion (ID) signature activity was generally low and more variable than SBS activity. In ED, ID19 (n=14) and ID23 (n=11) were the most recurrent indel signatures, whereas the replication-slippage signatures ID1 and ID2 were largely driven by the monocytic AML sample ED16 (Fig. 6B, E-F). Apart from ID2, all signatures increased with age in *TP53*-driven ED patients (Supplementary Fig. 15A-H).

**Fig. 6.**
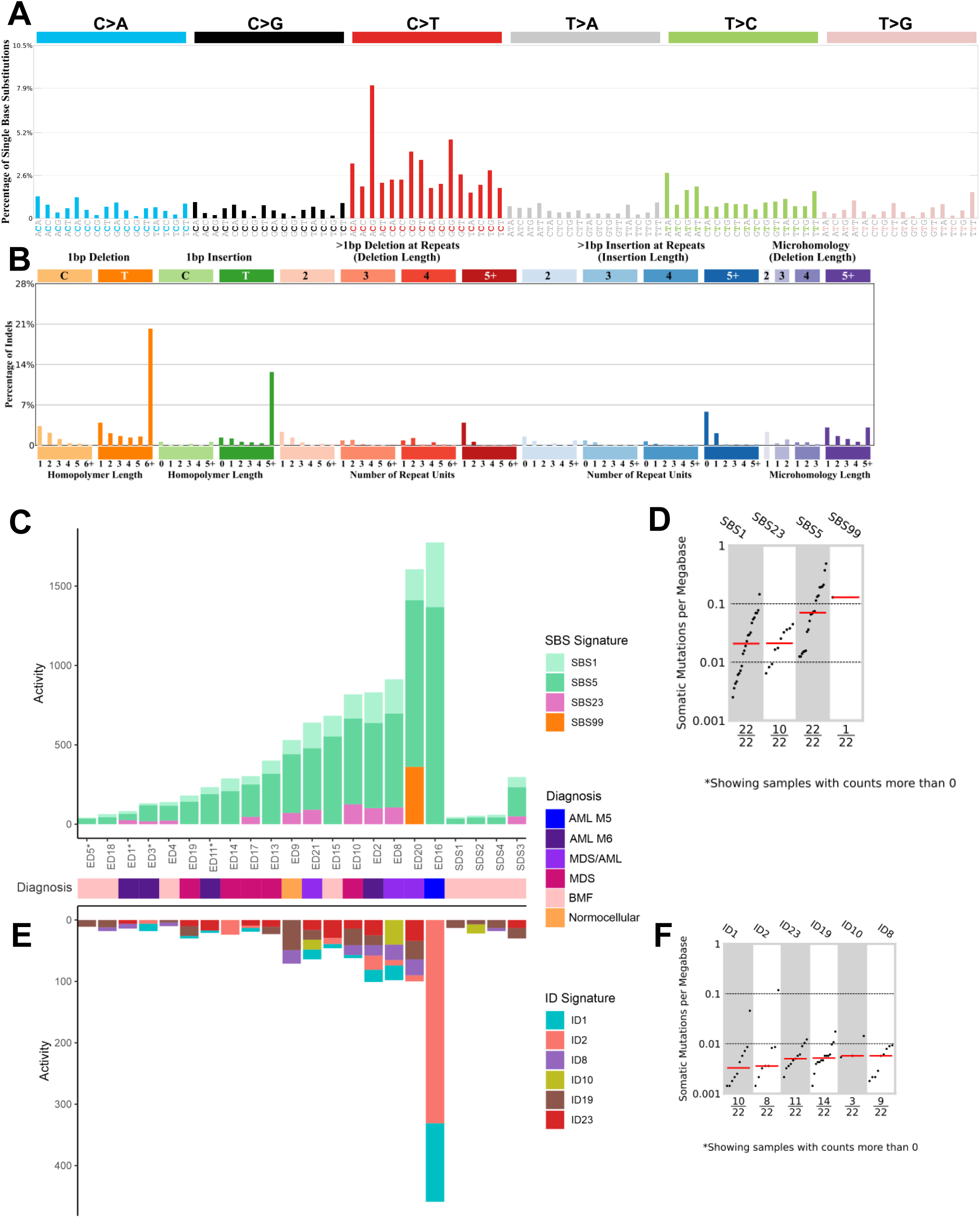
Mutational signatures in somatic ED and SDS samples. **A** SBS signature of the samples. **B** ID signature of the samples. **C** COSMIC SBS and **E** ID signature activities in each patient with WGS data. * Matched normal was FFPE-derived; signature activities may therefore be underestimated. **D** Mutation burden in each SBS signature per megabase. **F** Mutation burden in each ID signature per megabase. SBS, single-base substitution; ID, small insertion and deletion; WGS, whole-genome sequencing; FFPE, formalin-fixed paraffin-embedded.

### Kataegis accompanies chromosomal instability in ED-associated erythroid AML

Kataegis, defined as the clustered accumulation of somatic substitutions within restricted genomic regions, has been observed near structural rearrangements in cancer genomes^33,37^. In our cohort, kataegis was detected in 4/23 (17%) ED patients with evaluable WGS data and in 4/10 patients in the malignant phase (40%), all in MDS/AML or erythroid-type AML (Fig. 5, Table S11), but not in SDS samples.

In ED, kataegis events occurred on chromosomes with larger CNAs and often mapped within or adjacent to CNA regions and near breakpoints (Table S11). No recurrently affected gene was identified, suggesting that kataegis reflected localized genomic instability rather than recurrent targeting of specific drivers. Together, these findings indicate that kataegis is a feature of advanced, *TP53*-driven ED-associated erythroid malignancy.

## DISCUSSION

In this study, we define the somatic genomic evolution of ED across BMF to *TP53*-mutated AML. Our data provide evidence for ED representing potentially the most prominent preleukemic dependency on p53 pathway attenuation described in the literature. Although *ERCC6L2* is still absent from many routine cancer-predisposition gene panels, ED appears to occur at a frequency comparable to the better-established IBMFS such as SDS or Fanconi anemia (FA).

Clinically, ED progresses through three phases, translating into two genetic states (Fig. 7). During the BMF phase, ERCC6L2 deficiency creates hematopoietic stress and strong selective pressure on the p53 pathway but is not associated with broad clonally detectable genomic instability. The BMF phase typically persists for the first three decades of life, with only rare earlier progression to malignancy^2,16^. During malignant progression, complex rearrangements, kataegis, and chromothripsis-like features of p53-disrupted complex-karyotype myeloid malignancy are acquired. ED is distinguished by the recurrence and multiplicity of somatic *TP53* mutations: *TP53*-mutant clones are already detectable during BMF, before overt MDS or AML, and the burden of multiple *TP53*-mutant clones exceeds that of sporadic *TP53*-mutated myeloid neoplasia. Further, all *TP53*-negative ED patients were children, supporting *TP53*-mutant clonal selection as an early, characteristic feature of ED.

**Fig 7.**
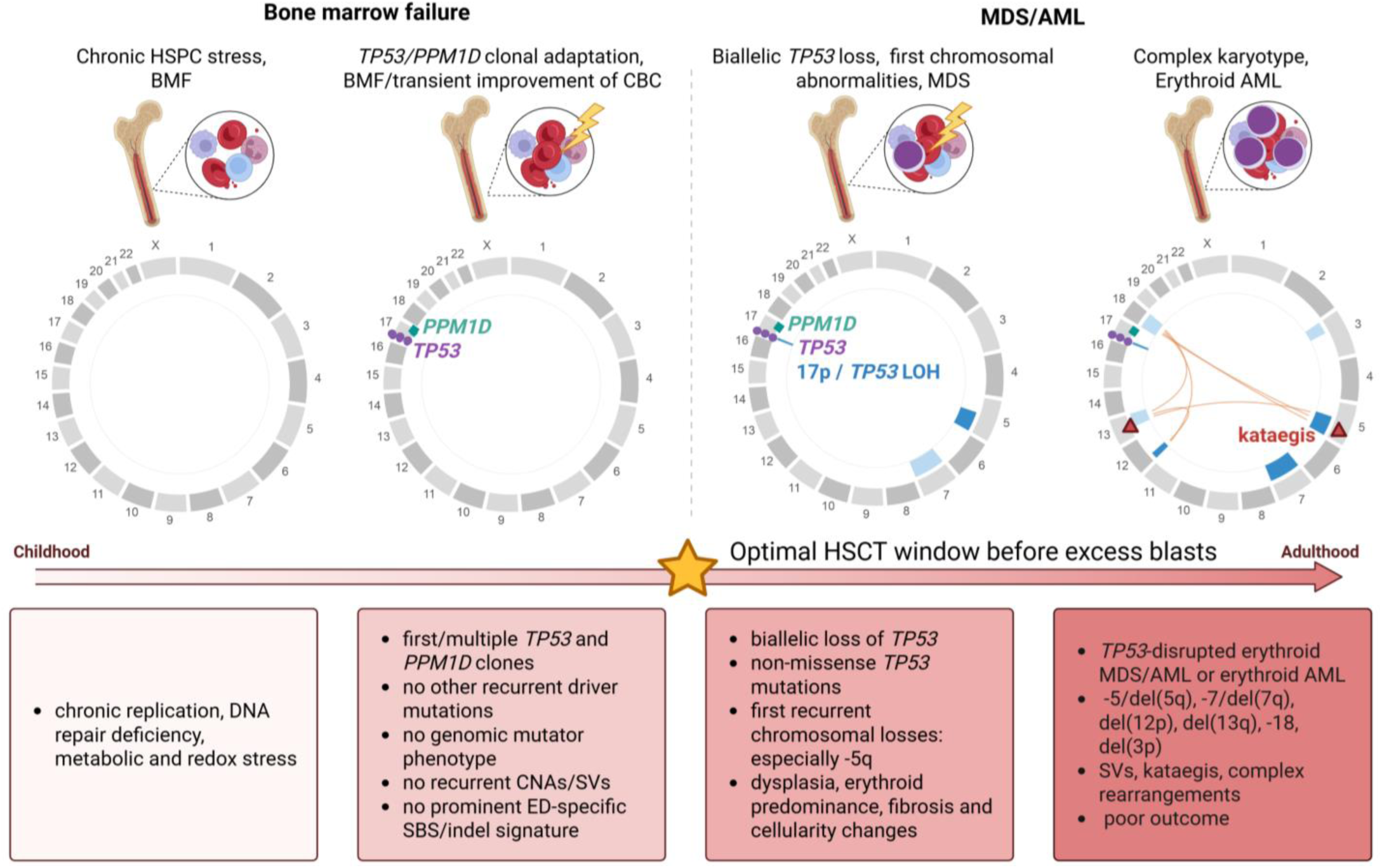
ERCC6L2 disease clonal evolution. Overview of stage-specific somatic evolution in ERCC6L2 disease from childhood BMF to adult erythroid-predominant MDS/AML. During the BMF phase, chronic hematopoietic stem and progenitor cell (HSPC) stress occurs in the absence of recurrent chromosomal abnormalities, structural variants, or a distinct genome-wide mutational signature. A subsequent adaptive phase is marked by emergence of *TP53* and/or *PPM1D*-mutant clones, with transient hematologic improvement but continued BMF. Progression to MDS is associated with biallelic *TP53* disruption, including non-missense *TP53* mutations, acquisition of the first recurrent chromosomal abnormalities, and erythroid predominance. Advanced disease is characterized by *TP53*-disrupted erythroid MDS/AML with complex karyotype, recurrent chromosomal losses, structural complexity, and kataegis. AML, acute myeloid leukemia; BMF, bone marrow failure; CBC, complete blood count; CNA, copy-number alteration; HSPC, hematopoietic stem and progenitor cell; HSCT, hematopoietic stem cell transplantation; MDS, myelodysplastic syndrome; SBS, single-base substitution; SV, structural variant.

Most of the early *TP53* alterations were missense mutations, consistent with selection for altered or dominant-negative p53 function^38^. In contrast, non-missense *TP53* mutations were observed only in ED patients who had already transformed to MDS/AML or progressed shortly after clone emergence. Biallelic *TP53* inactivation through distinct mechanisms appears to be a critical step in progression to AML^39^. Although multiple *TP53* mutations do not necessarily indicate biallelic inactivation, non-missense mutations may act as functional second hits and often occurred without *TP53* CNA/CN-LOH. While the prognostic impact of truncating *TP53* mutations remains unsettled^40,41,38,42^, recent work suggests that different *TP53* mutation classes may have distinct roles in initiation and progression of myeloid neoplasia. Inactivating mutations are enriched in AML with myelodysplasia-related changes and transcriptionally active *TP53* mutations are more common in MDS^43^, consistent with our findings in ED. The stage-dependent shift from missense-dominated selection during BMF to non-missense, biallelic disruption at transformation, provides a longitudinal human correlate to the functional distinctions recognized in *TP53*-mutated myeloid neoplasia and may be the most generalizable feature of ED. Non-missense *TP53* mutations in ED, particularly when arising alongside pre-existing *TP53*-mutant clones, appear to be high-risk events that should prompt evaluation for HSCT before progression to MDS with excess blasts or overt AML.

In our cohort, the only other recurrently mutated driver gene in ED was *PPM1D*. PPM1D negatively regulates the DNA damage response and p53 signaling, and hotspot mutations increase its stability and activity, enabling cells to tolerate genotoxic stress, as reflected by the recurrent selection of *PPM1D*-mutant clones in therapy-related clonal hematopoiesis, telomere-biology disorders, and aging ^30,44–49^. Although their malignant potential in treatment-naïve patients remains uncertain ^50,51^, *PPM1D* mutations likely confer a selective advantage in ERCC6L2-deficient hematopoiesis without correcting the underlying defect.

The restricted somatic landscape distinguishes ED from other IBMFS. In SDS, clonal evolution can proceed through compensatory *EIF6* mutations or del(20q), partially rescuing the ribosome defect^29,52^. In FA, chr1q gain and *MDM4* overexpression similarly attenuate p53 signaling and improve HSPC fitness^53^. In contrast, we found no comparable compensatory route in ED, which instead was dominated by *TP53* and *PPM1D* alterations.

Recent functional work provides insight into the strong selection for *TP53*-mutant clones in ED. Ercc6l2-deficient hematopoietic stem and progenitor cells experience replication stress and DNA damage that activate *Trp53*, causing cell-cycle arrest, apoptosis, and impaired fitness; *Trp53* inactivation partially rescues the hematopoietic defect but not the replication stress, identifying p53 activation as a major mediator of BMF^14^. ERCC6L2 is also required for faithful repair of staggered-end double-strand breaks, providing a mechanistic basis for the DNA-repair vulnerability^12^. Our previous work demonstrated that *TP53* selection occurs in a BM already shaped by erythroid dysfunction: ED cells showed upregulated stress programs, and during progression *TP53*-mutant cells acquired aberrant erythroid priming rather than returning to normal hematopoiesis^3^. Thus, ED progresses from ERCC6L2-driven stress to *TP53*-mediated somatic rescue and, finally, *TP53*-disrupted complex-karyotype malignancy.

Malignant ED samples were characterized by recurrent chromosomal abnormalities and complex genome architecture overlapping that of *TP53*-mutated myeloid malignancies. Particularly, chromosome 5 and 7 alterations appeared to be early events in several patients, consistent with −5/del(5q) and −7/del(7q) as recurrent early events in *TP53*-mutated complex-karyotype MDS/AML^54,55^. Together with later 12p, 18 and 3p losses, some of which diminished following treatment, and the erythroid-lineage enrichment of 5/7/12p aberrations, these data support branched chromosomal evolution underlying erythroid-predominant malignant progression.

Several recurrent lesions also affected genes previously linked to *TP53*-mutated or complex-karyotype myeloid disease. Recurrent 12p deletions encompassed ETV6 and CDKN1B (reported in 45% of *TP53*-mutated AML/MDS^6^ and 49% of complex-karyotype AML^56^), 13q loss likely acted through RB1 (described in *TP53*-mutated erythroid leukemias and linked to poor survival^57^), and 5q loss may reflect combined haploinsufficiency of driver and anti-driver genes rather than a single target^58^. Losses of 5q and 3p also overlap with recently described *TP53*-mutated AML involving 3p/5q ribosomal-protein-gene losses, creating an erythroid-associated ribosomopathy-like phenotype^59^. The recurrent co-occurrence of 3p/*BAP1* deletion with *TP53* inactivation parallels the *BAP1/TP53* erythroleukemia model^60^, in which *Bap1* loss cooperates with *Trp53* deficiency to transform erythroid-primed progenitors; its dependence on BCL2L1/BCL-xL is consistent with *BCL2L1* overexpression in ED erythroid cells^3^. Although speculative, these parallels suggest that selected chromosomal losses in *TP53*-disrupted ED contribute functionally to erythroid malignant progression.

Like the structural and copy-number landscape, the SBS and indel signatures revealed no strong ED-specific genome-wide mutational process; ERCC6L2 deficiency is reflected more by *TP53/PPM1D* selection, CNAs, SVs, and complex chromosomal instability than by a unique SNV or indel signature. Given the established link between chromothripsis and *TP53* alterations in complex-karyotype AML^61^, the kataegis and chromothripsis-like features restricted to malignant, *TP53*-disrupted erythroid-type disease likely reflect *TP53*-driven complex-karyotype evolution rather than ERCC6L2 deficiency itself. Overall, the combination of *PPM1D*-mutant clonal hematopoiesis, recurrent p53 pathway lesions, and complex cytogenetic evolution places ED close to therapy-related, *TP53*-mutant erythroid AML, with ERCC6L2 deficiency acting as an endogenous genotoxic stressor that selects for p53-pathway attenuation.

Our study is limited by a the modest cohort size, which reflects the rarity and relatively recent recognition of ED. Larger longitudinal series are difficult to assemble, and sample collection is further constrained by the clinical urgency of treatment at progression. Our SDS comparison cohort was also limited and lacked leukemic-phase samples. Thus, the ED–SDS comparisons are most informative for the BMF phase and early clonal evolution.

Together, our findings suggest that the defining somatic feature of ED is not an early genome-wide mutator phenotype, but early and recurrent selection for *TP53*-pathway attenuation. Malignant progression is marked by transition to biallelic *TP53* disruption through loss of heterozygosity or a non-missense second hit, accompanied by recurrent chromosomal losses, chromothripsis-like structural complexity, kataegis, and an erythroid-predominant phenotype. Thus, ERCC6L2 deficiency creates a constrained evolutionary landscape in which *TP53* and *PPM1D* are the dominant routes of clonal selection, whereas overt genomic instability becomes evident after biallelic *TP53* inactivation and malignant transformation. By resolving the trajectory in a germline setting of exceptionally strong selective pressure, ED provides a window into how intense, stage-dependent selection on the *TP53* pathway shapes the transition from BMF to *TP53*-mutant leukemia more broadly.

## AUTHOR CONTRIBUTIONS

**Suvi P. M. Douglas**: Conceptualization; data curation; investigation; formal analysis; methodology; software; visualization; writing —original draft, writing —review and editing. **Ilse Kaaja**: Conceptualization; visualization; investigation; writing—original draft; writing—review and editing. **Ina Ikonen**: Data curation; software; visualization. **Jessica Koski**: Data curation; formal analysis; software; visualization. **Laura Langohr**: Data curation; investigation; formal analysis; software; visualization, writing —review and editing. **Marja Hakkarainen**: Data curation; resources. **Lotta Katainen**: Data curation. **Gisela Barbany**: Resources. **Eva Hellström-Lindberg**: Resources. **Kirsi Jahnukainen**; Resources. **Sakari Kakko**: Resources. **Timo Siitonen**: Resources. **Riitta Niinimäki**: Resources. **Esa Pitkänen**: Conceptualization; methodology; supervision; writing—review and editing. **Ulla Wartiovaara**-**Kautto**: Conceptualization; supervision; funding acquisition; resources; writing —original draft, writing—review and editing. **Outi Kilpivaara**: Conceptualization; supervision; funding acquisition; writing —original draft, writing—review and editing.

## DATA AVAILABILITY STATEMENT

The data generated in this study derive from patients with a rare disease and are therefore individually identifiable. In accordance with the terms of the participants’ written informed consent and the approval of the Coordinating Ethics Committee of the Helsinki University Hospital, and under the EU General Data Protection Regulation, these data cannot be deposited in a public repository. Summary-level data, including variant calls, allele frequencies, and clonal composition supporting the findings of this study, are provided in the manuscript and its supplementary files.

## ETHICS STATEMENT

The study was conducted in accordance with the Declaration of Helsinki. The study has been approved by Helsinki University Central Hospital ethics review committee (#206/13/03/03/2016, amendment 2023, and HRUHLAB2). All samples from living individuals are derived after written informed consent.

## ACKNOWLEDGEMENTS

We would like to thank Pihla Siipola, Tuulia Räisänen and Kirsi Kvist-Mäkelä for technical assistance, Sadiksha Adhikari for locating data, and Siiri Reinikka for expertise in SV interpretation and valuable comments on figures. Sequencing was performed by FIMM Genomics NGS Sequencing unit at the University of Helsinki supported by HiLIFE and Biocenter Finland and HUSLAB Laboratory of Genetics HUS Diagnostic Center, Helsinki University Hospital. The authors wish to acknowledge the IT Center for Science (CSC), Finland, and the Institute for Molecular Medicine Technology Center (FIMM TC) for generous computational resources. The study benefited from samples from Northern Finland Biobank Borealis, Oulu, Finland (https://oys.fi/en/front-page/for-researchers/biobank-borealis-of-northern-finland/).

ChatGPT (OpenAI) and Claude (Anthropic) were used to modify or generate scripts and revise text. The authors carefully reviewed all AI-assisted edits and take full responsibility for the content of the manuscript.

