## Supplementary information for "Intense *TP53* pathway selection drives clonal evolution from bone marrow failure to leukemia in ERCC6L2 disease"

*Douglas et al.*

### Supplementary information

#### SUPPLEMENTARY METHODS

##### Clinical NGS panel sequencing and karyotyping

Somatic mutations of most patients were previously determined for diagnostic purposes in the Laboratory of Genetics, Diagnostic Center, Helsinki University Hospital using an in-house-developed NGS myeloid panel as described previously[1] or an updated version of the gene panel (Table S6.). The target regions were covered on average by >5000 reads. Mutations with a variant allele fraction (VAF) of ≥2% were considered. For samples collected in Oulu, the somatic mutations were determined for diagnostic purposes using Sophia Genetics extended myeloid custom version C library kit. Generally, mutations with a VAF of ≥5% were considered, but for *TP53* also smaller VAFs were reported. BM karyotyping and/or fluorescence in situ hybridization (FISH) was performed at the center of care of the patient as part of the diagnostic workup.

##### Samples for whole-genome sequencing

Patient skin biopsies were obtained in conjunction with diagnostic BM sampling. Germline DNA was extracted from cultured skin fibroblasts (if available), skin biopsy, buccal swab or archived formalin-fixed paraffin-embedded non-malignant tissue for three deceased patients (Table S1) using DNeasy Blood and Tissue Kit (Qiagen, Hilden, Germany) according to protocol. For one patient, germline DNA was extracted from a buccal swab with DNeasy Blood & Tissue kit (Qiagen). For three deceased patients, only archived Formalin-Fixed Paraffin-Embedded (FFPE) non-malignant tissue samples were available, and germline DNA was extracted using standard phenol-chloroform method.

Somatic DNA was extracted from fresh BM aspirate (or peripheral blood sample if not available) with Nucleospin Blood Kit (Macherey-Nagel, Düren, Germany) according to protocol. For one patient only archived methanol-fixed BM was available. DNA quantification was done with Qubit DNA assay (Thermo Fisher Scientific, Waltham, MA, USA). Genomic DNA integrity was examined with Tapestation (Agilent Technologies, Santa Clara,CA, USA).

**Whole genome sequencing and secondary data analysis**

Libraries were constructed at Functional Genomics Unit Helsinki with NEBNext Ultra II FS DNA (New England Biolabs, Ipswich, MA, USA) or Illumina DNA PCR-Free (Illumina, San Diego, CA) kits according to protocols. Whole genome sequencing (WGS) was conducted on Novaseq 6000 (Illumina, San Diego, CA) 2x150bp by Institute for Molecular Medicine Finland (FIMM) or the Laboratory of Genetics, Diagnostic Center, Helsinki University Hospital. Data quality control was done with Fastqc (v0.12.1) (https://www.bioinformatics.babraham.ac.uk/projects/fastqc/). Secondary data analysis was done on the Illumina Dragen Bio-IT platform (v.4.4.6) at FIMM. Reads were aligned to hg38. Average coverage was 66x for somatic samples and 34x for germline samples.

**Somatic variant filtering**

Variants called by Dragen were filtered and annotated with Baseplayer[2] (v. 1.0.2). Variants with minor allele frequency (MAF) of <0.001 in gnomAD genomes (v4.1)[3], at least four variant reads, minimum coverage of 10x, minimum variant allele frequency (VAF) of 5% and somatic quality score (SQ) ≥20 were considered. Only variants not in GRCh38 unified blacklist[4] were considered to avoid problematic regions of the genome. Variants of likely germline origin (variant seen in multiple patients without matching normal or reported in gnomAD genomes and VAF 0.4-0.6 or >0.9) or variant MAF >0.001 in gnomAD Finns were discarded. Variants were visually inspected with Integrative genomics viewer (IGV) (v.2.14.0), and suspicious variants (variants in poorly mapped reads, repeat regions and/or with low VAF, few supporting reads in only one direction) were discarded. *TP53* and *PPM1D* were also manually inspected for variants with VAF<5%. Variant descriptions were confirmed with VariantValidator[5,6] (v. 3.0.2).

For mutational signatures and clustered mutation analyses, only samples with a matching normal were used and only canonical chromosomes (chr1-22, X, Y) were included. To analyze only high-confidence variants, we used the following parameters for filtering: SQ≥20, VAF ≥5%, minimum coverage ≥10x, at least four variant reads, no variant reads in matching normal sample, variant not in GRCh38 unified blacklist regions or in panel of normals constructed from all germline WGS samples.

**Structural variant filtering**

To include only high-confidence structural variants (SVs), we set the requirements as follows: at least five alternative-supporting fragments in the tumor, at least three tumor split reads, at most one alternative-supporting fragment in the matched germline sample, and a tumor-to-germline variant fragment ratio of at least five. All SVs were visually inspected using IGV and suspicious variants (variants seen in other germline samples, with suspicion of germline origin or very little evidence for a true variant) were discarded.

**Data visualization**

Oncoplot was made using R package ComplexHeatmaps[7] (v.2.26.1). *TP53* and *PPM1D* mutation lollipop visualization was made with ProteinPaint[8]. CNA heatmaps constructed based on Dragen segmentation data were generated with R (v.4.5.3) package GenVisR[9]. B-allele frequencies (BAF) for each sample were plotted with karyoploteR[10].

For Circos[11] (v.0.69.8) visualization, copy-number and B-allele frequency segmentation files generated by Dragen were converted into Circos-compatible tracks. Only canonical chromosomes were retained. For copy-number visualization, Segment_Mean values were centered by subtracting 1, yielding 0 for copy-neutral segments and positive or negative values for gains and losses, respectively; segments with absolute centered values ≤0.1 were omitted to reduce noise. For allelic imbalance visualization, BAF segments were represented using the BAF_SLM_STATE annotation, excluding segments with undefined values, and only states 1–9 were plotted. All high-confidence SVs, regardless of driver status, were plotted. For kataegis, clustered single-nucleotide variants were grouped by chromosome and groupNumber. For each kataegis cluster, the mean genomic position of all variants within the cluster was calculated and used as the cluster midpoint.

### RESULTS

**Mutational signatures**

Most single base substitutions (SBS) in ED and SDS samples were attributed to the clock-like signatures SBS1 and SBS5, which are widely observed in normal and malignant tissues and increase with age (Fig. 6A,C-D). No ED-specific signature was found, but the SNV burden in ED reflects endogenous age-associated mutagenesis previously described in normal hematopoietic stem and progenitor cells and in pediatric AML/HSPC comparisons[12,13].

SBS1 is associated with spontaneous deamination of 5-methylcytosine and behaves as a mitotic/age-related clock, whereas SBS5 is also clock-like but has less certain etiology.[12,13] Consistent with this, both SBS1 and SBS5 increased with age in ED patients (Supplementary Fig. 15A-B). A minor contribution from SBS23 was detected in ten patients, predominantly in ED. SBS23 has unknown etiology but has been reported to show transcriptional strand bias compatible with transcription-coupled nucleotide excision repair.[12,13] In ED, SBS23 contribution also increased with age (Supplementary Fig. 15C). One patient (ED20) showed a contribution from SBS99, a signature experimentally associated with melphalan exposure, but in the absence of known exposure, this finding was interpreted cautiously as possible signature bleeding or misassignment rather than true melphalan-associated mutagenesis. No significant strand-bias was detected for any signature. Doublet base substitutions (DBS) were rare, preventing reliable comparison with known COSMIC DBS signatures.

Small insertions and deletion (ID) signature activity was generally low across ED and SDS samples and more variable than SBS activity. Across ED samples with detectable ID activity, ID19 (n=14) and ID23 (n=11) were the most recurrent signatures, while the replication-slippage signatures ID1 and ID2 were seen in multiple samples but were largely driven by the monocytic AML (ED16) as a clear outlier in our ED patient set (Fig. 6E). ID19 has unknown etiology and has been reported in hematologic cancers and sarcomas[14], while ID23 is associated with aristolochic acid exposure[15], making its assignment in our cohort tentative in the absence of known exposure or matching SBS22/DBS20 activity. Low-level contributions from ID8, linked to double-strand break repair by non-homologous end joining, and ID10 of unknown etiology, were also observed in individual samples (Fig.

6E-F)[16]. Apart from ID2, most ID signatures increased with age (Supplementary Fig. 15D-H).
