## Supplementary figures for "Intense *TP53* pathway selection drives clonal evolution from bone marrow failure to leukemia in ERCC6L2 disease"

*Douglas et al.*

Supplementary Figures

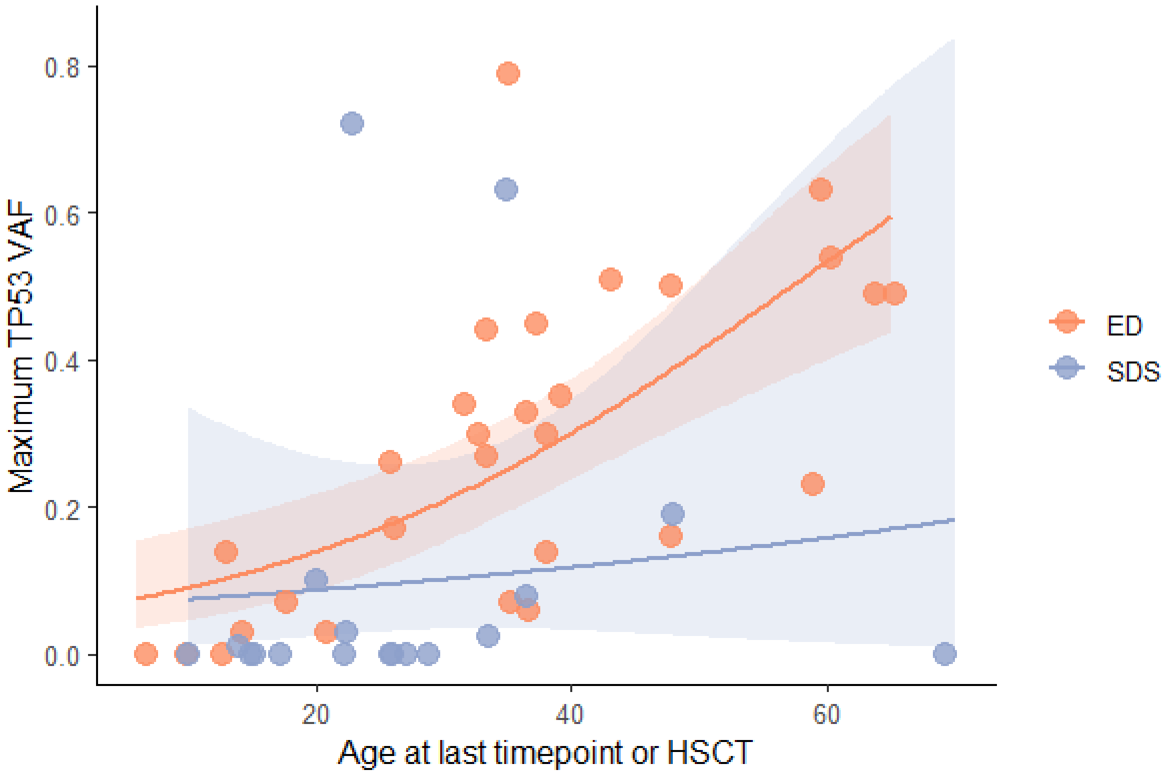


**Supplementary Figure 1. Age-associated expansion of *TP53*-mutant clones in ED and SDS.** The variant allele frequency (VAF) of the largest *TP53* mutation clone per patient plotted against age at latest follow-up or last pre-treatment/pre-HSCT timepoint. Lines show fractional logistic regression fits, with shaded areas indicating 95% confidence intervals.

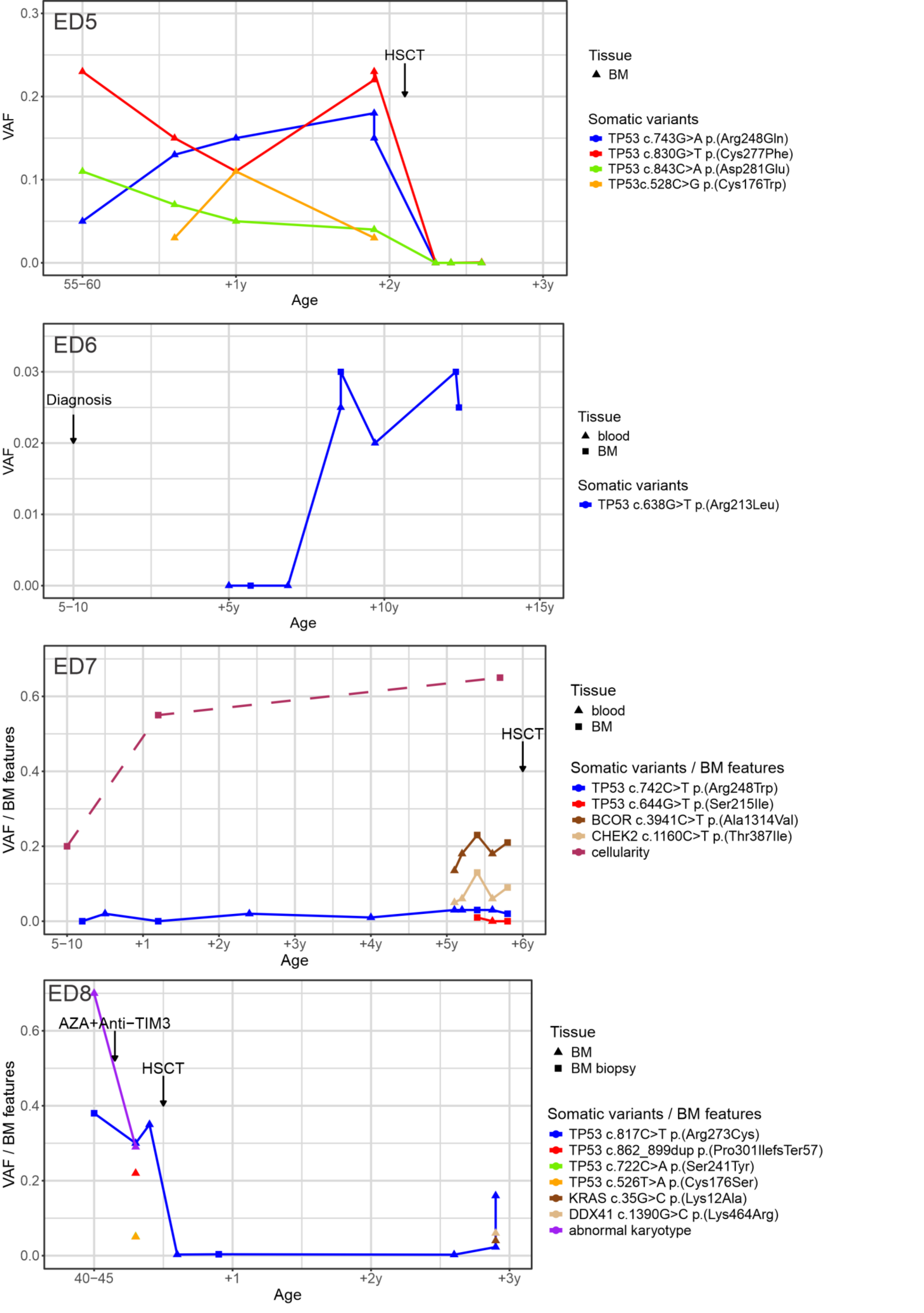


**Supplementary Figure 2. Changes in *TP53* and other somatic potential driver mutation variant allele fraction (VAF) over time in ED patients ED5, ED6, ED7 and ED8.** Bone marrow features (cellularity, myeloid:erythroid (M:E) ratio, percentage of erythropoiesis (EPO) if available) and treatments from patients with four or more timepoints.

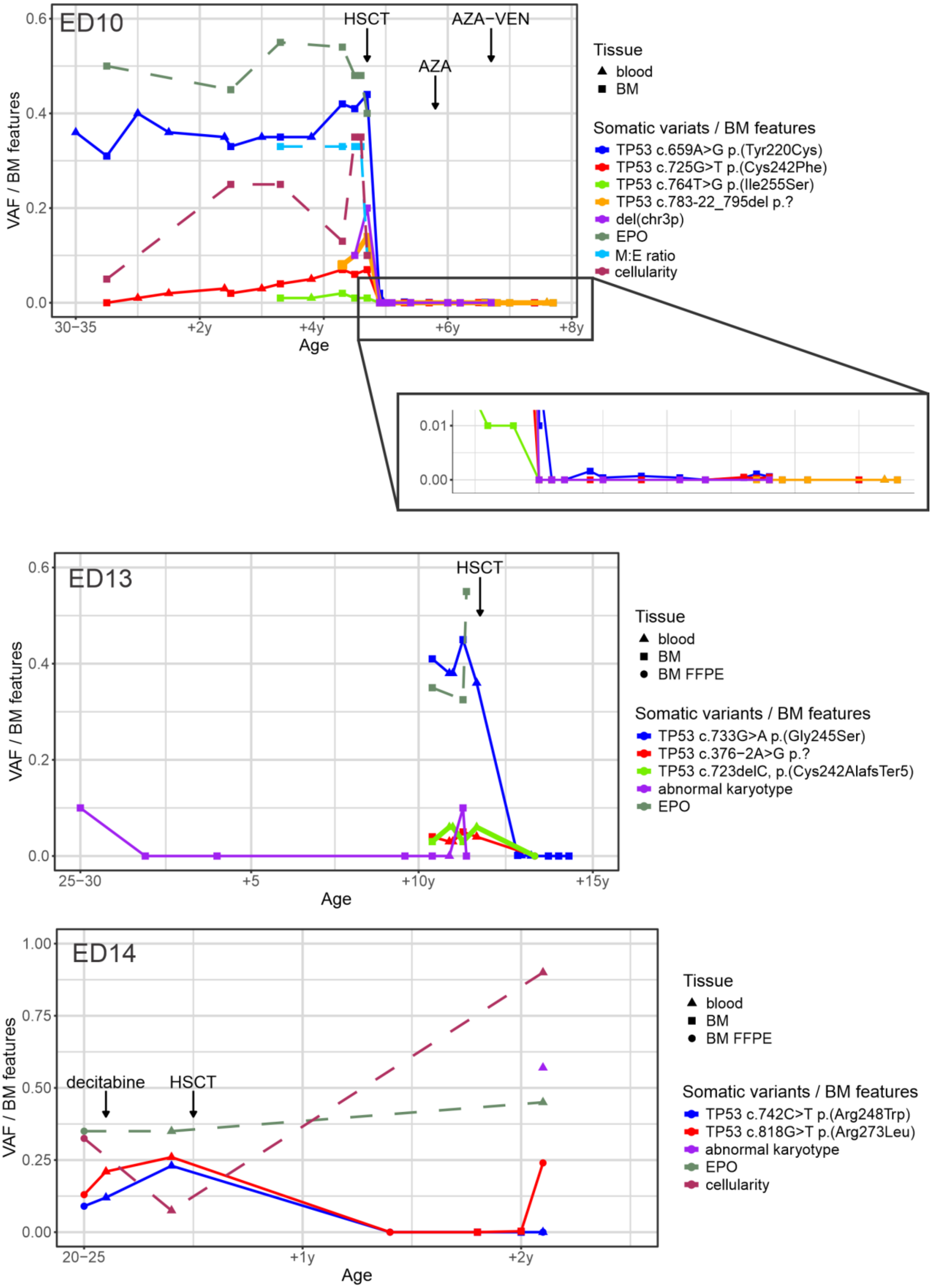


**Supplementary Figure 3. Changes in TP53 and other somatic potential driver mutation variant allele fraction (VAF) over time in ED patients ED10, ED13 and ED14.** ED10 shows slight clone increase after HSCT. Bone marrow features (cellularity, myeloid:erythroid (M:E) ratio, percentage of erythropoiesis (EPO) if available) and treatments from patients with four or more timepoints.

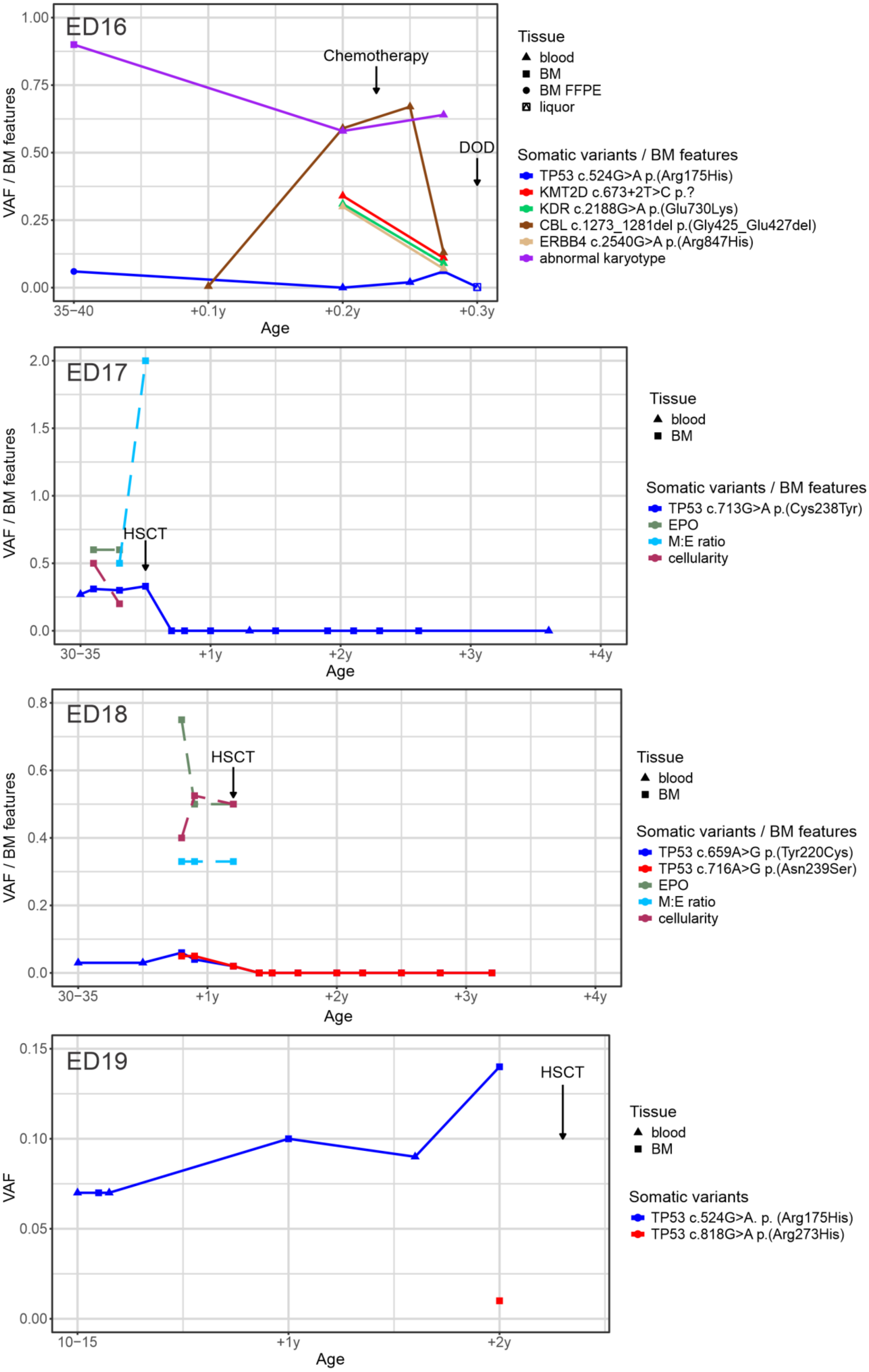


**Supplementary Figure 4. Changes in TP53 and other somatic potential driver mutation variant allele fraction (VAF) over time in ED patients ED16, ED17, ED18 and ED19**. Bone marrow features (cellularity, myeloid:erythroid (M:E) ratio, percentage of erythropoiesis (EPO) if available) and treatments from patients with four or more timepoints.

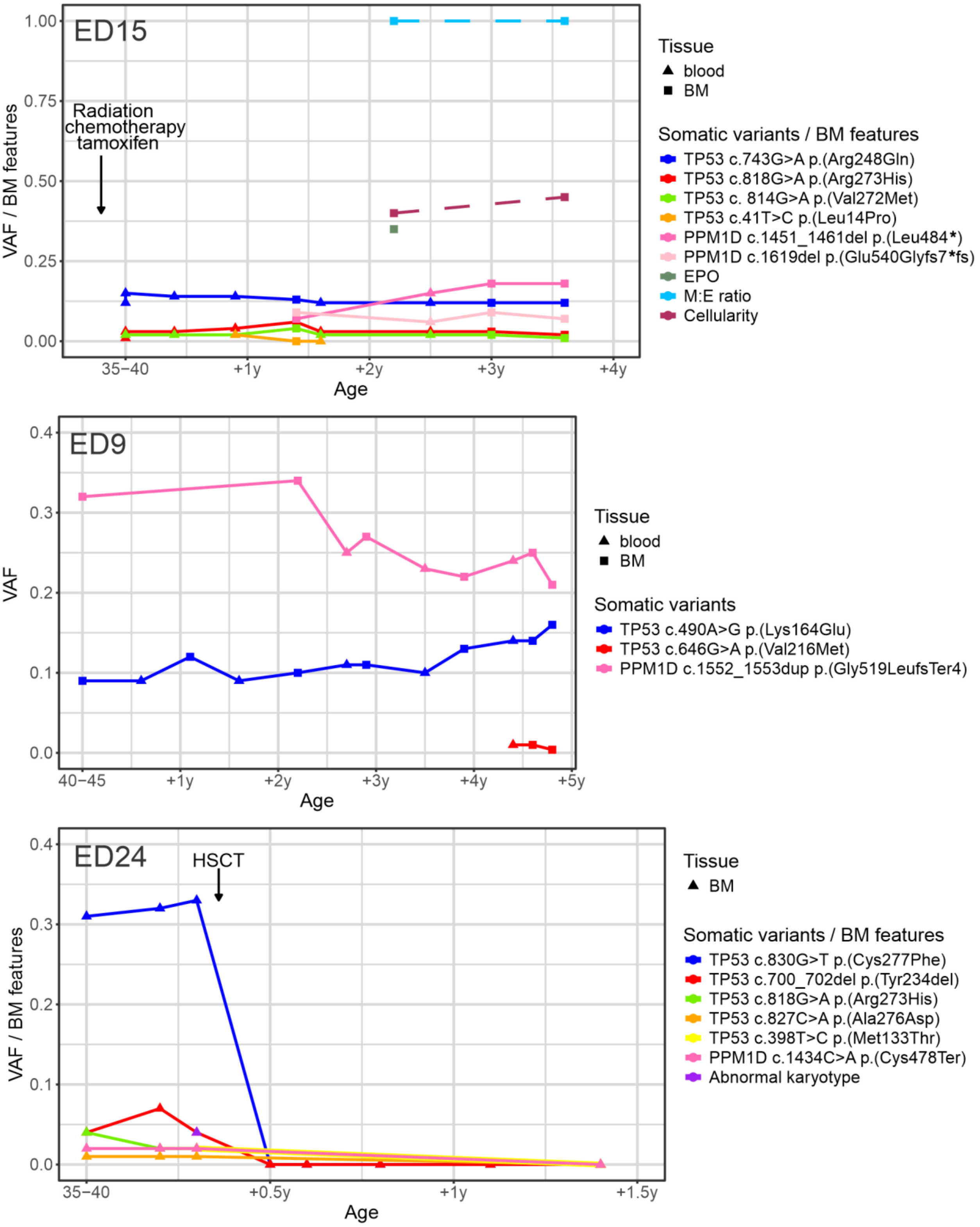


**Supplementary Figure 5. Changes in TP53 and other somatic potential driver mutation variant allele fraction (VAF) over time in PPM1D-mutated ED patients ED15, ED9 and ED24.** Bone marrow features (cellularity, myeloid:erythroid (M:E) ratio, percentage of erythropoiesis (EPO) if available) and treatments from patients with four or more timepoints.

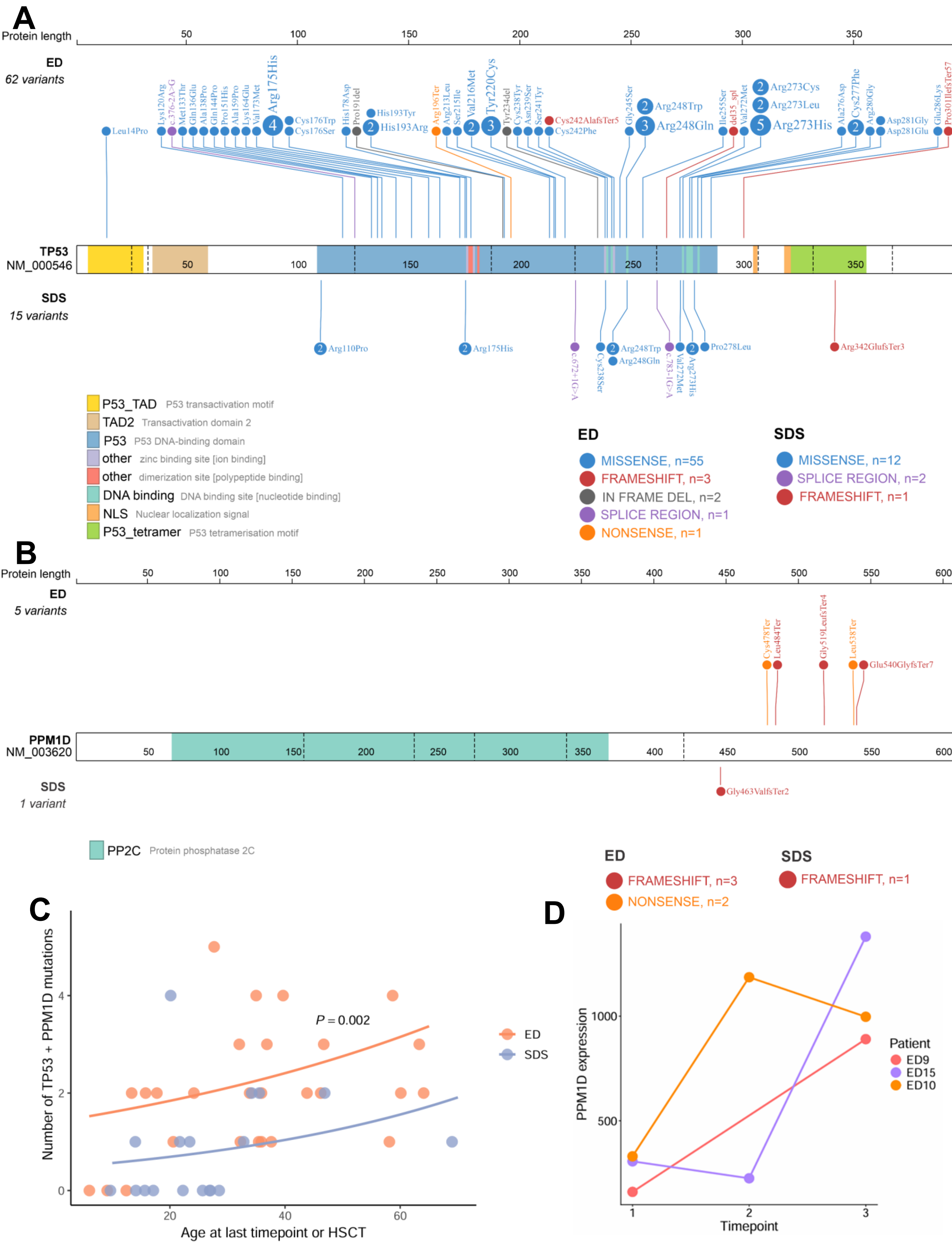

**Supplementary Figure 6. *TP53* pathway mutations in ED and SDS patients. A** *TP53* mutations in ED and SDS patients are mostly in the DNA-binding domain of p53. **B** *PPM1D* mutations in ED and SDS are frameshift mutations affecting the last exon. **C** The number of *TP53* pathway mutations (*TP53* + *PPM1D*) per patient plotted against age at latest follow-up or last pre-treatment/pre-HSCT timepoint. Lines indicate fitted values from a Poisson regression model including age and condition. The P value denotes the age-adjusted difference in combined *TP53* and *PPM1D* mutation count between ED and SDS. **D** *PPM1D* expression in serial scRNAseq samples from ED bone marrow.

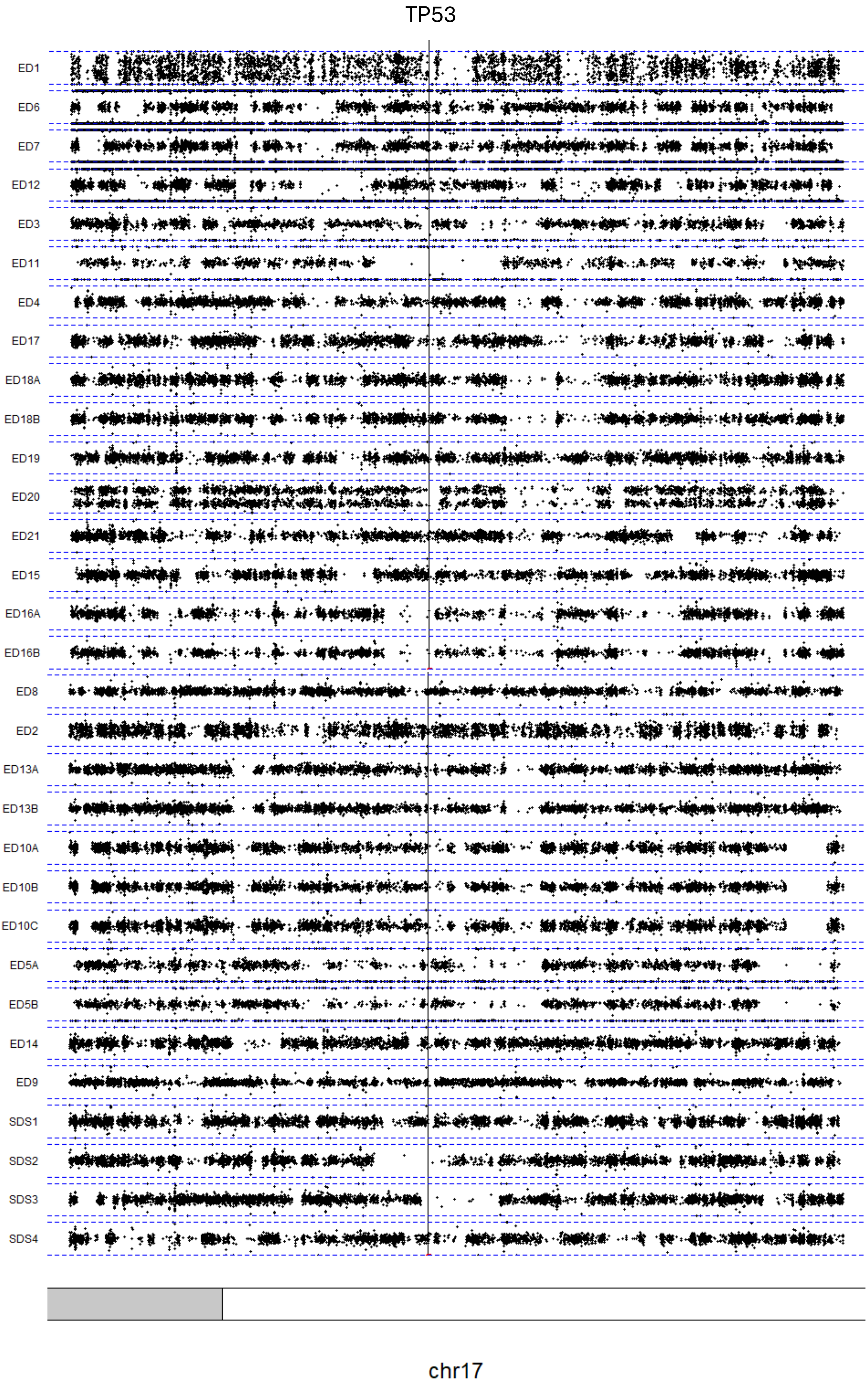


**Supplementary Figure 7. B-allele frequency (BAF) on chromosome 17p around TP53 locus for each sample.** Deviation of BAF from 0.5 indicates loss of heterozygosity.

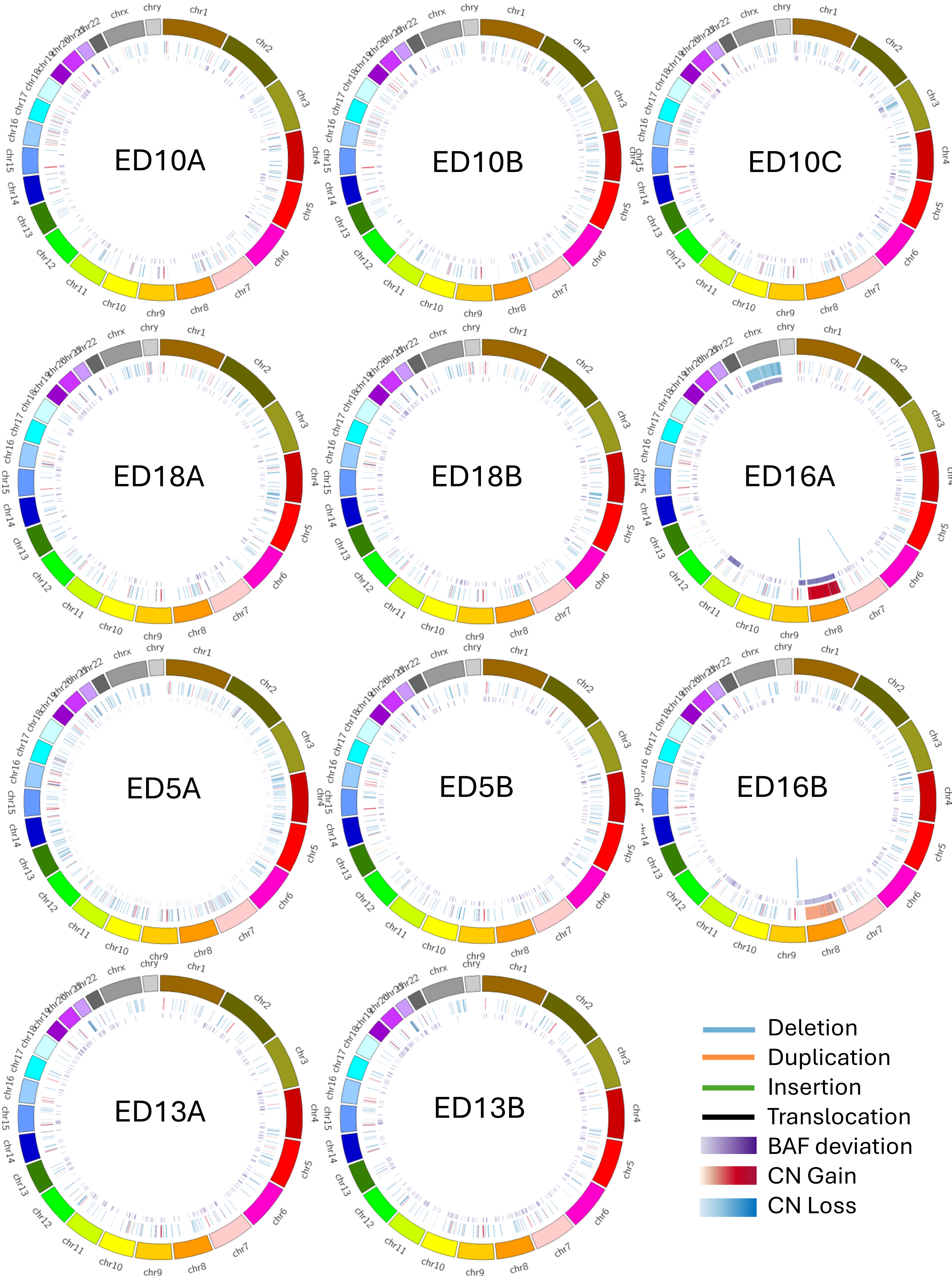


**Supplementary Figure 8. Copy number (CN) and structural variants in WGS samples with multiple timepoints from segmentation data.** Red corresponds to copy number gain, blue corresponds to copy number loss, purple corresponds to B-allele frequency (BAF) deviation from 0.5 - brighter color indicating stronger deviation from normal copy number.

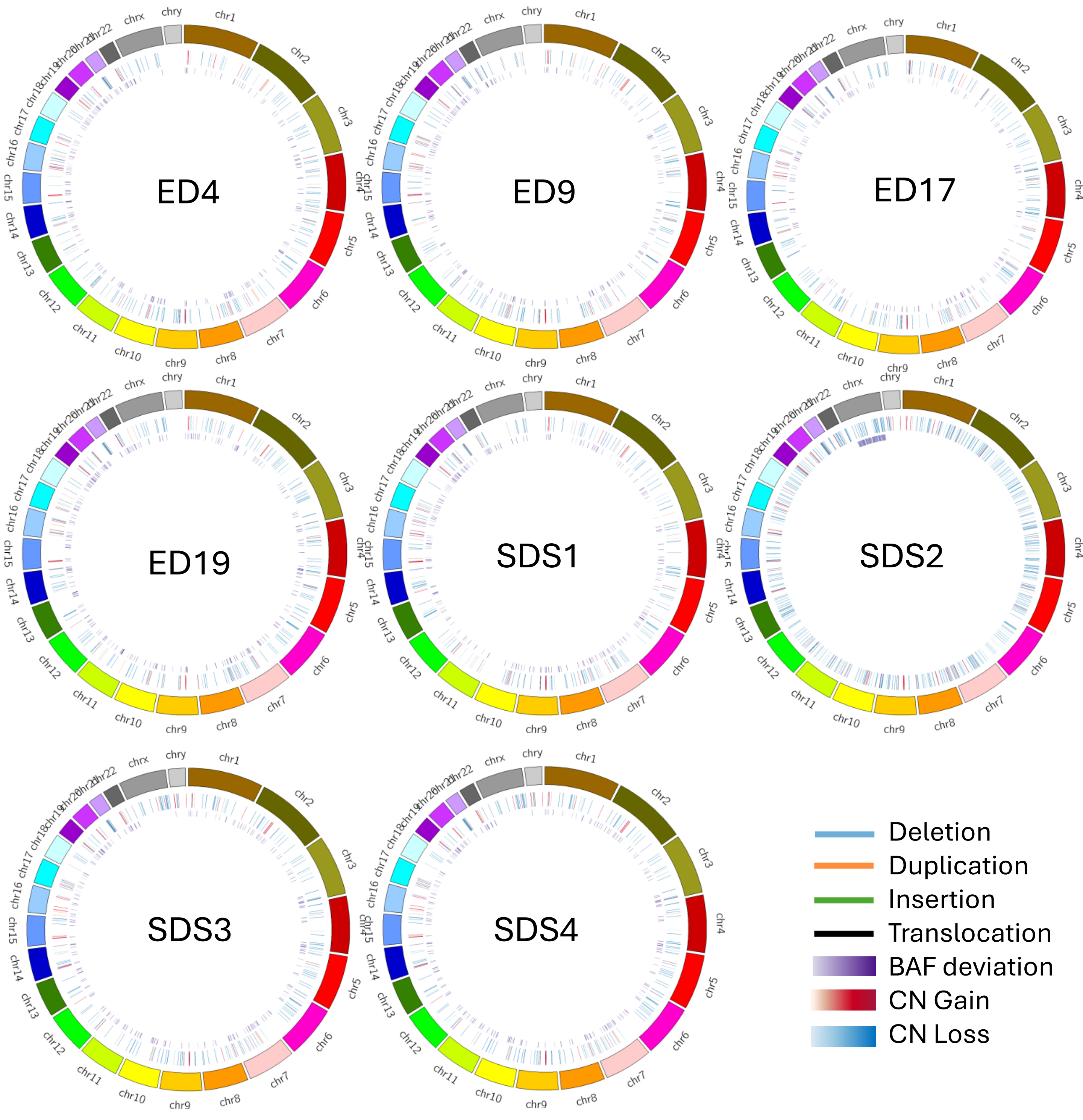


**Supplementary Figure 9. Copy number (CN) and structural variants in WGS samples from ED patients with no chromosomal abnormalities and single timepoint and all SDS patients from segmentation data.** Red corresponds to copy number gain, blue corresponds to copy number loss, purple corresponds to B-allele frequency (BAF) deviation from 0.5 - brighter color meaning stronger deviation from normal copy number.

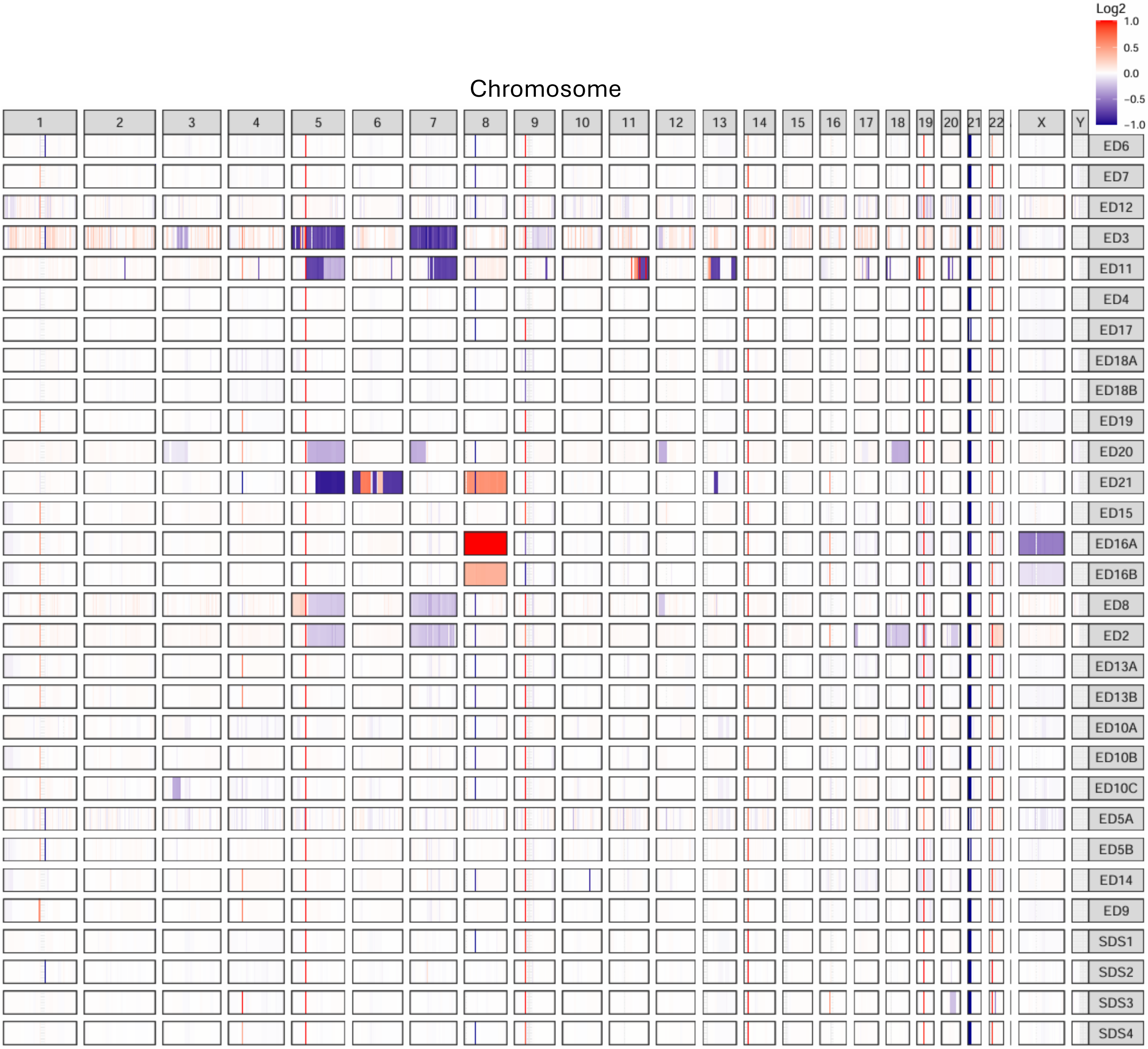


**Supplementary Figure 10. Copy number in WGS samples from DRAGEN segmentation data.** Red corresponds to copy number gain, blue corresponds to copy number loss, brighter color indicates stronger deviation from normal copy number (log2 scale).

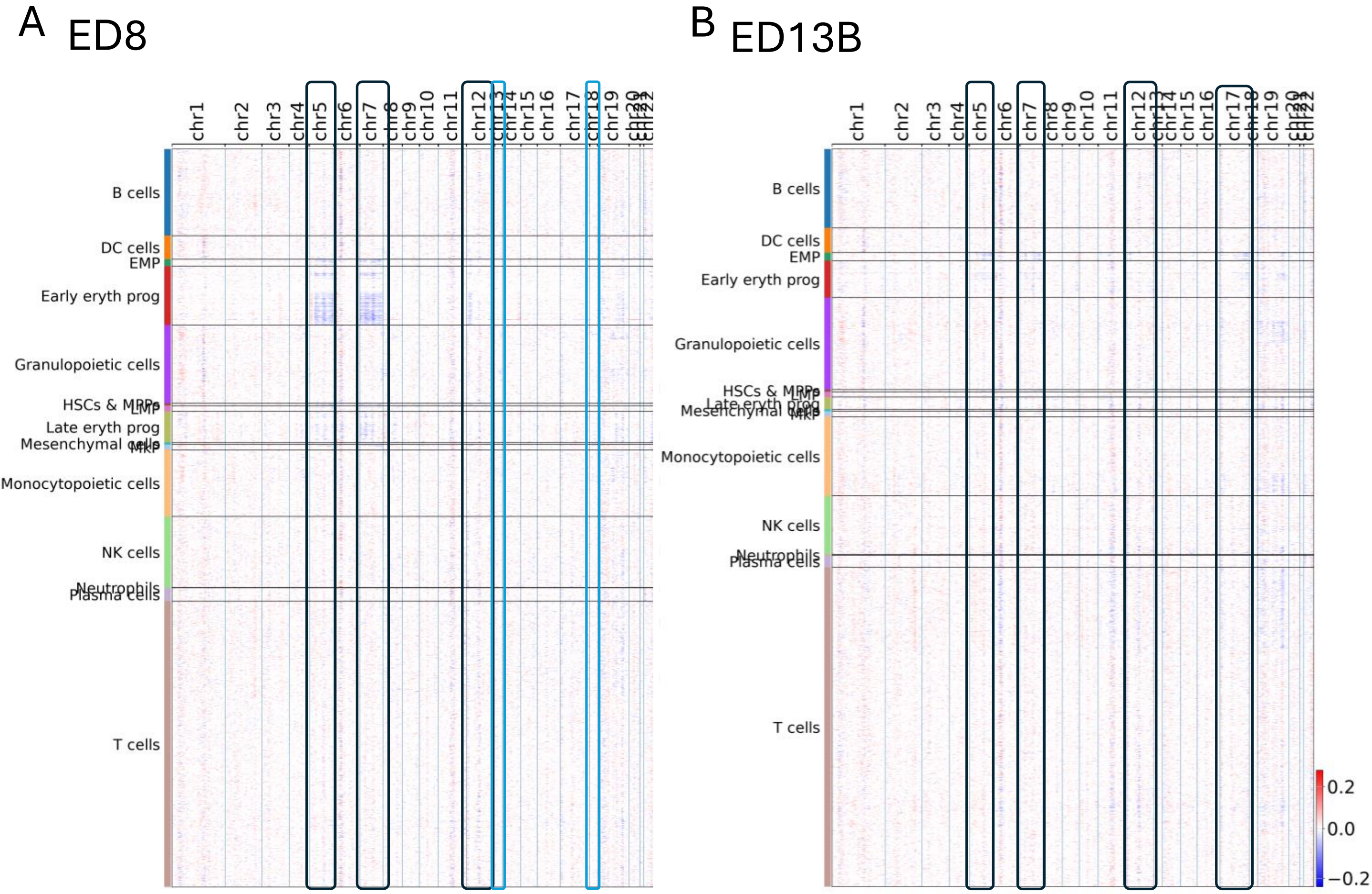


**Supplementary Figure 11. Copy number in different cell types inferred from single-cell RNA sequencing data in patients ED8 and ED13B.** Boxes highlight chromosomes with alterations. Black box shows alterations seen in erythroid lineage cells and blue boxes show alterations on chromosomes 13 and 18 in myeloid cell types, affected by treatment and of which only traces are seen in ED8.

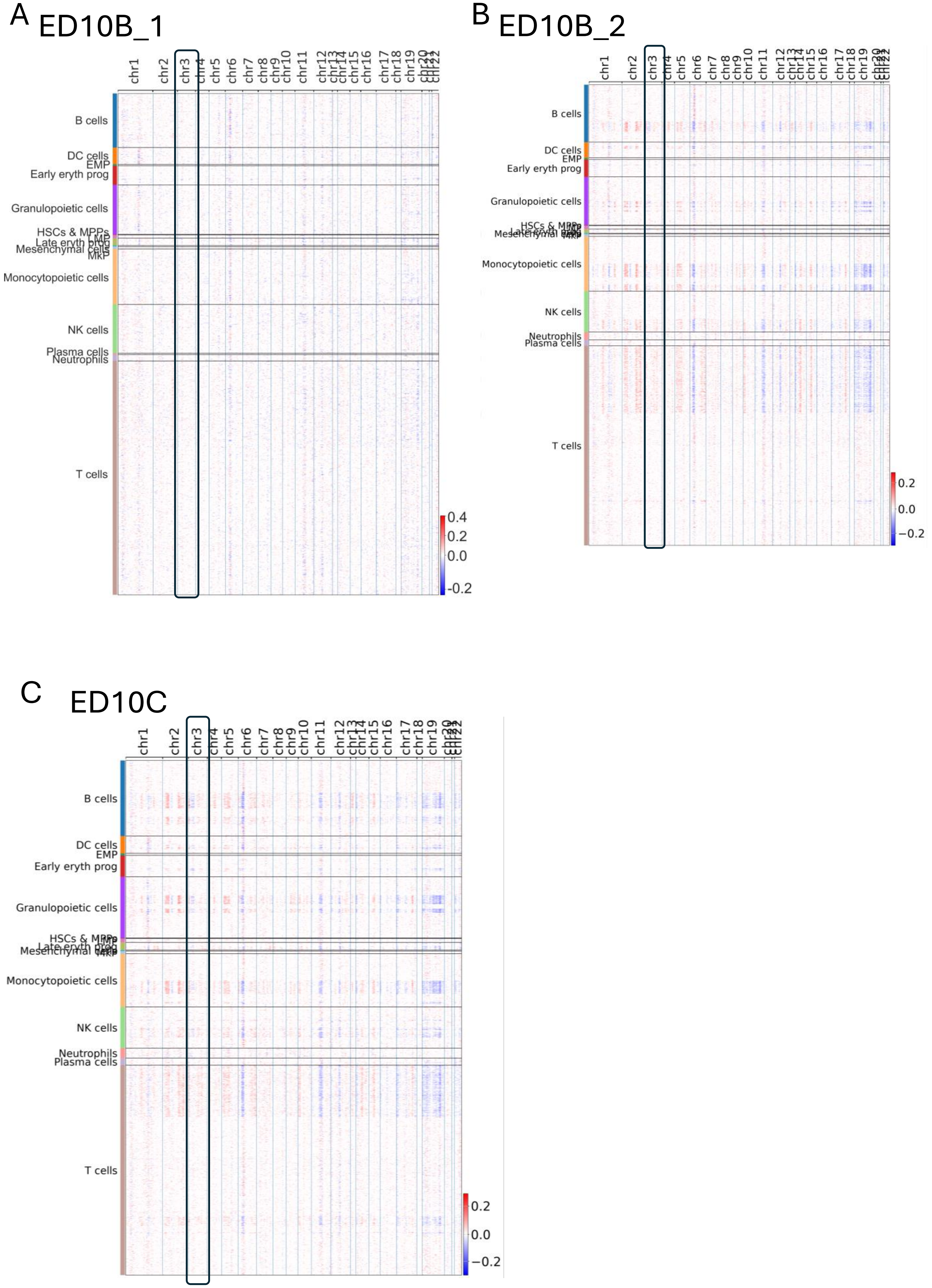


**Supplementary Figure 12. Copy number in different cell types inferred from single-cell RNA sequencing data in ED10 over time.** Boxes highlight chromosome 3 where 3p deletion occurred. Only traces are visible at the first timepoint (**A**), whereas the deletion is evident at subsequent timepoints (**B-C**).

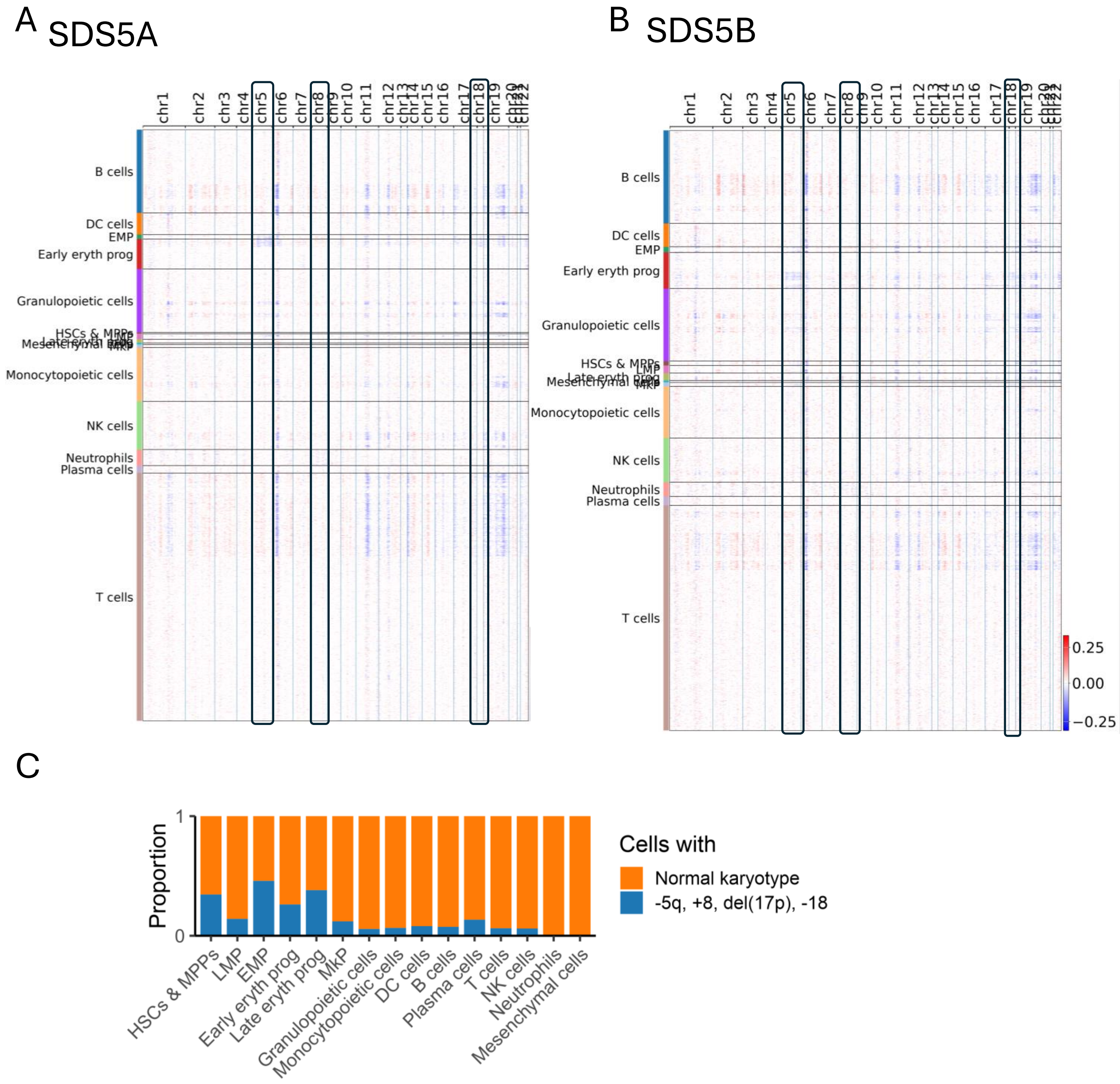


**Supplementary Figure 13. Copy number in different cell types inferred from single-cell RNA sequencing data in SDS5 at two timepoints. A-B** Boxes highlight chromosomes with copy-number alterations. **C** Proportions of cells with abnormal and normal karyotypes by cell type.

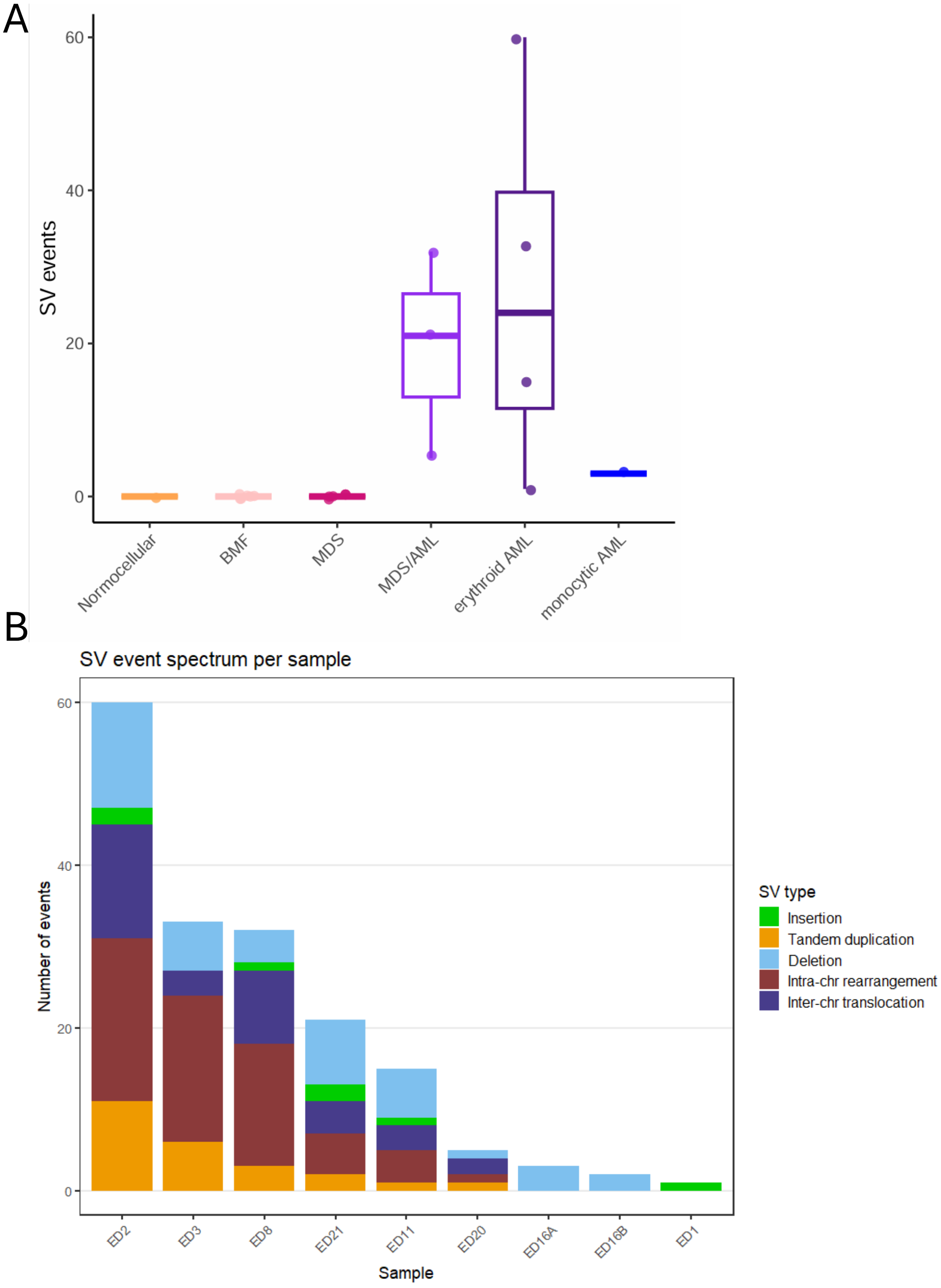


**Supplementary Figure 14. Number of structural variant (SV) events in ED patients. A** SV events by diagnosis. **B** SV events by type per sample. AML, acute myeloid leukemia; BMF, bone marrow failure; MDS, myelodysplastic syndrome; MDS/AML, MDS with excess blasts.

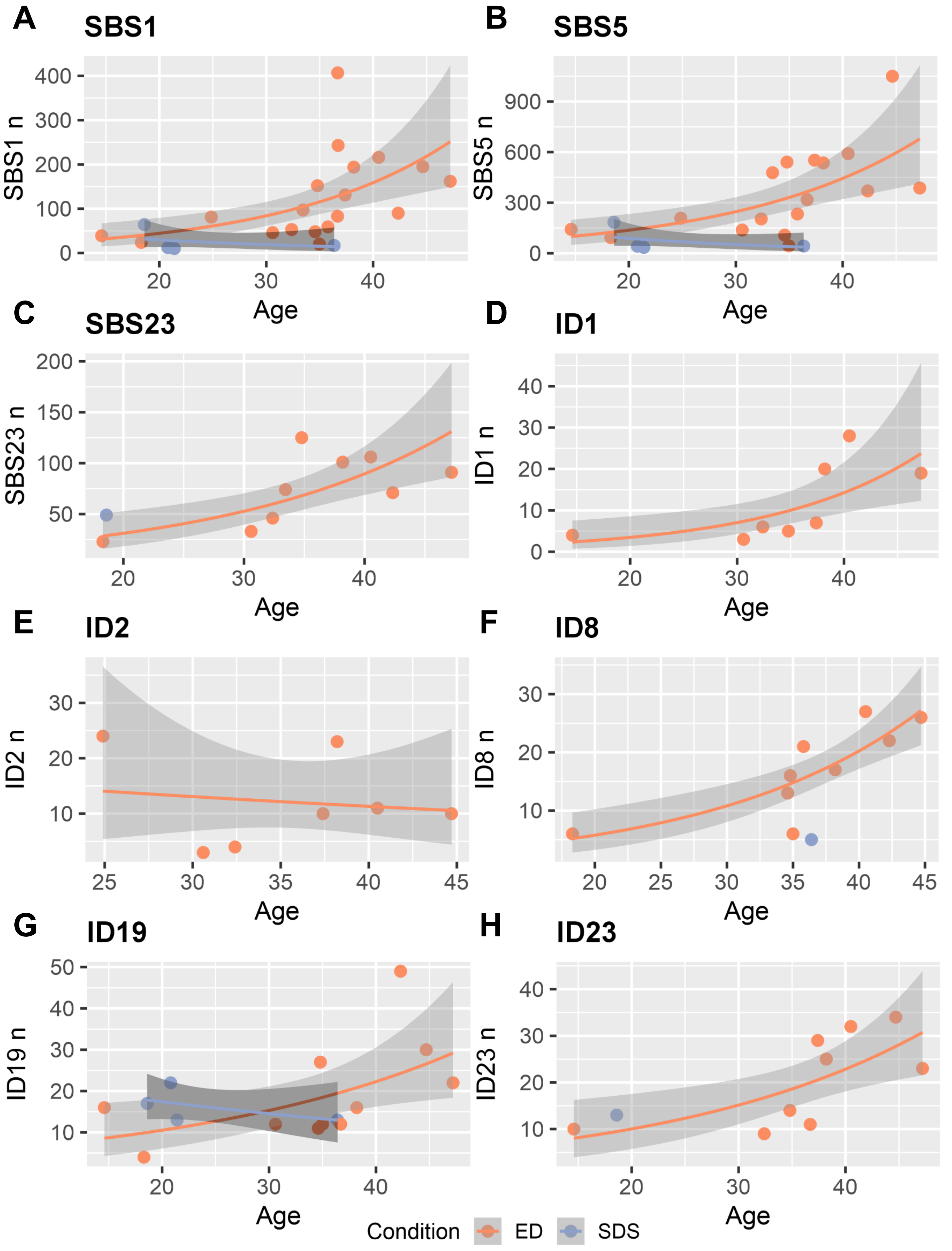


**Supplementary Figure 15. All signatures except ID2 increase with age in TP53-driven ED patients**. Only samples with activity in signature are shown, samples with FFPE normal not shown due to underestimation of activities. FFPE, formalin-fixed paraffin-embedded; ID, small insertion and deletion signature; SBS, single-base substitution.
